# Shared Clinical and Immunologic Features of mRNA Vaccines: Preliminary Results from a Comparative Clinical Study

**DOI:** 10.1101/2024.11.26.24318005

**Authors:** Carlos Fierro, Nelia Sanchez-Crespo, Daniel Makrinos, Weijie Zhang, Yanbo Sun, Poonam Rohilla, Bethany Girard, Abidemi Adeniji, Anthony DiPiazza, Robert Paris

## Abstract

**Introduction:** Clinical trials do not typically assess underlying molecular mechanisms of vaccine immunogenicity or reactogenicity. We evaluated the reactogenicity and immunogenicity of 4 mRNA vaccines and potential contributing mechanisms and identified shared and unique clinical and immunologic features.

**Methods:** This ongoing open-label, phase 1 trial randomized healthy adults (18-75 years) to receive a single dose of mRNA-1273.222 (bivalent COVID-19), mRNA-1345 (RSV), mRNA-1010 (influenza), and FLUAD (active influenza comparator) or 2 or 3 doses of mRNA-1647 (CMV). The primary objective was to assess the safety and reactogenicity of each study vaccine, with humoral immunogenicity (neutralizing antibody [nAb] responses) as the secondary objective. This interim analysis reports safety and reactogenicity in all study vaccines and humoral immunogenicity in single-dose vaccines (mRNA-1273.222, mRNA-1345, mRNA-1010, and FLUAD). Exploratory objectives included antigen-specific T-cell responses after single-dose mRNA-1345 or mRNA-1273.222, and soluble mediators of inflammation and innate immunity following vaccination in single-dose vaccine groups and two doses of mRNA-1647.

**Results:** At the interim analysis data cutoff (February 1, 2023), 302 participants received 1 dose of the study vaccines. Reactogenicity exhibited a consistent trend across vaccine groups; most solicited local and systemic adverse reactions within 7 days were mild or moderate in severity. There were no deaths or serious, severe, or treatment-related adverse events leading to study discontinuation. At Day 29, nAb titers against vaccine-specific antigens increased 2- to 8-fold versus baseline for all single-dose vaccine groups. In an exploratory analysis, mRNA-1273.222 and mRNA-1345 induced antigen-specific Th1-biased CD4^+^ and CD8^+^ T-cell responses at Day 29. The cytokine response analysis showed increased levels of IFN-γ, IL-6, IL-2Ra, CXCL9, IP-10, MCP-2, and MIP-1β on Day 2 following vaccination, with generally greater increases observed with mRNA vaccines versus FLUAD. Regardless of age and across mRNA vaccine groups, peak serum levels of IL-1Ra and MCP-1/MCP-2 on Day 2 weakly correlated with systemic reactogenicity scores (correlation coefficient range: 0.15-0.27).

**Conclusions:** The 4 mRNA vaccines had acceptable reactogenicity, demonstrated changes in serum biomarkers of innate immune activation, and were immunogenic. This suggests that the observed reactogenicity of mRNA vaccines may be related to shared features of the mRNA platform (LNP platform).

**Clinical Trial Registration:** ClinicalTrials.gov NCT05397223

## 1 Introduction

Vaccines that elicit broadly protective and long-lasting immunity are needed to prevent severe outcomes from many pathogens, including respiratory and latent viruses. Compared with traditional vaccines (eg, DNA-based and live or attenuated viral vaccines), mRNA vaccines have several notable advantages, including rapid and scalable manufacturing, and ease of adaption to new antigen designs [1,2]. The acceptable safety profile, immunogenicity, and effectiveness of the mRNA-based COVID-19 vaccine, mRNA-1273 (Spikevax; Moderna, Inc., Cambridge, MA, USA) [3–7], has demonstrated the utility of such a platform for novel vaccine development. Additionally, a novel respiratory syncytial virus (RSV) mRNA vaccine (mRNA-1345) has been shown to be efficacious against RSV-lower respiratory tract disease and RSV acute respiratory disease in adults aged ≥60 years, with no evidence of safety concerns in a phase 3 trial [8]; mRNA-1345 has been approved in the United States for the prevention of RSV in adults ≥60 years [9]. Indeed, several other mRNA-based vaccine candidates are in clinical development, including those against cytomegalovirus (CMV; mRNA-1647), influenza (mRNA-1010), as well as combination vaccines (eg, mRNA-1083 against influenza and COVID-19) [10–12].

The underlying molecular mechanisms of vaccine immunogenicity or adverse reactions (reactogenicity) are infrequently assessed in vaccine clinical trials, which typically assess only vaccine-specific adaptive immune responses from peripheral blood samples collected at infrequent intervals. A comprehensive systems biology approach has been used primarily to define potential mechanistic correlates of protective immunity for several different vaccine platforms including attenuated virus, viral vector, adjuvanted recombinant protein, and conjugate vaccines, which have shared determinants of vaccine immunogenicity [13–18].

Although the mechanisms by which an mRNA-vaccine stimulate the immune system have been previously studied [19], it is still unknown whether there are any common or unique immune signatures that may be associated with or predictive of vaccine reactogenicity and immunogenicity for mRNA-based vaccines. Developing a better understanding of differences in the magnitude or patterns of change in biomarkers through in-depth characterization of innate and adaptive immunity induced by different mRNA-encoded antigens may help to identify characteristics common to mRNA vaccines as a class and lead to further improvements in mRNA vaccine efficacy and tolerability.

To this end, we initiated a phase 1, open-label, randomized, 2-part study to evaluate the safety, reactogenicity, and immunogenicity of modified mRNA vaccines in healthy adults and to investigate the potential mechanisms of both reactogenicity (solicited adverse reactions [ARs]) and immunogenicity using a systems biology approach. The vaccines included in this study have all progressed beyond phase 1 testing and include the COVID-19 mRNA-1273.222 (Original + BA.4/BA.5 bivalent, 1:1 mixed; approved for use as SpikeVax 2022/2023) and RSV mRNA-1345 vaccines, the seasonal quadrivalent influenza mRNA-1010 vaccine, and the CMV mRNA-1647 vaccine. mRNA-1273.222, mRNA-1345, and mRNA-1010 were evaluated for their ability to boost pre-existing vaccine- or virus-specific responses following a single vaccine dose. mRNA-1647 was evaluated as a 2- or 3-dose series in participants who were either CMV-seropositive or CMV-seronegative at baseline. The adjuvanted, enhanced seasonal quadrivalent influenza vaccine, FLUAD, served as a control for mRNA-1010. Age-related differences in vaccine responses were also explored. Herein, we report early results of the safety and immunogenicity, including immediate post-vaccination soluble biomarkers of inflammation, of each study vaccine.

## 2 Materials and Methods

### 2.1 Trial design and participants

This ongoing, open-label, randomized, 2-part, phase 1 trial (NCT05397223) evaluated the safety, reactogenicity, and immunogenicity of mRNA-1273.222, mRNA-1345, mRNA-1647, mRNA-1010, and FLUAD (active influenza comparator) in healthy adults (18-75 years). The mRNA-1273.222, mRNA-1345, mRNA-1010, and FLUAD arms enrolled participants in 2 age cohorts (n=∼30 each; 18-49 and 50-75 years); the 2 mRNA-1647 arms enrolled 30 participants (18-49 years) each (**Figure 1**).

**Figure 1.**
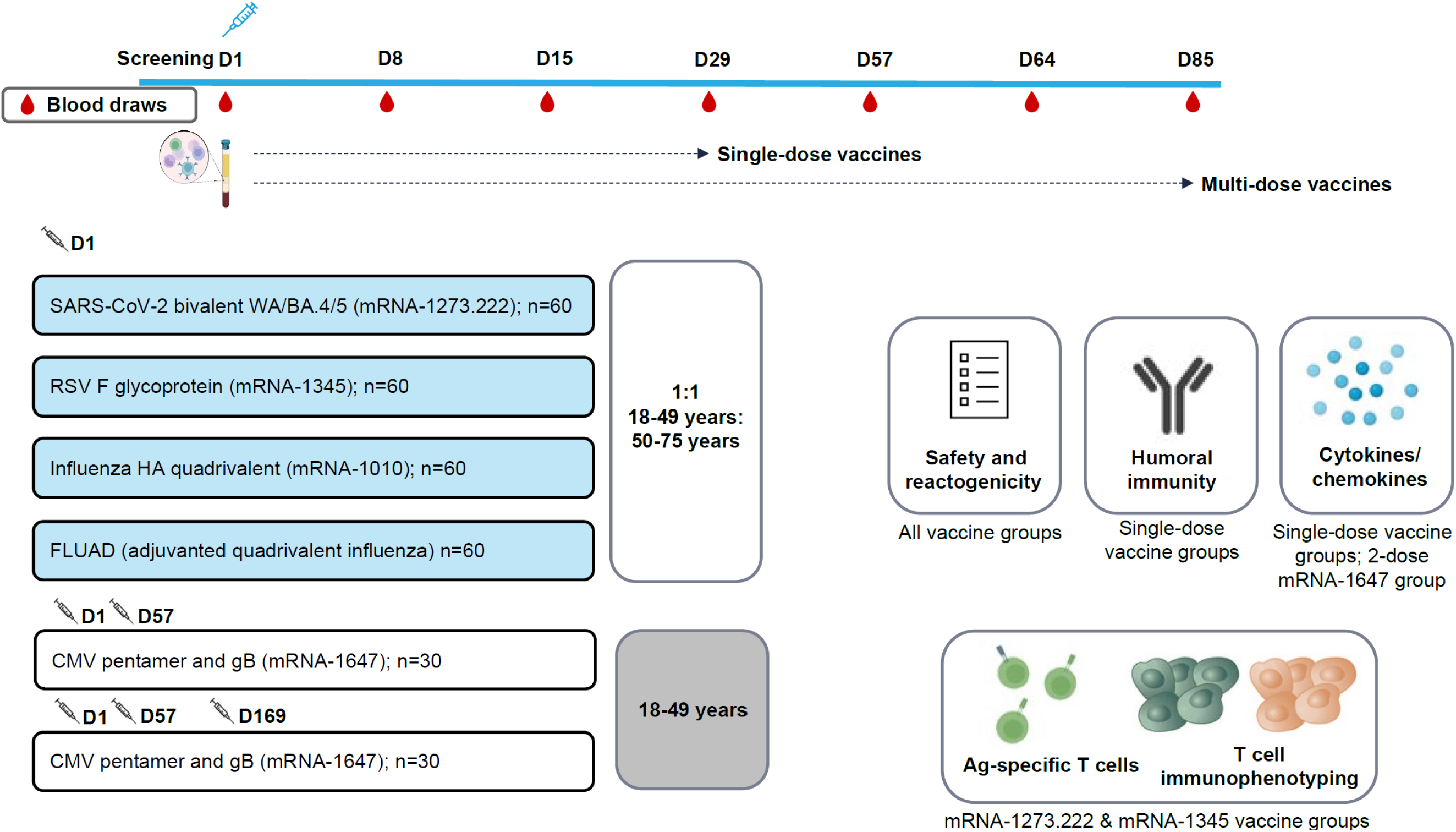
Study schema. Ag, antigen; BA.4/BA.5, Omicron-BA.4/BA.5; CMV, cytomegalovirus; D, Day; gB, glycoprotein B; FLUAD = adjuvanted (MF59), inactivated, quadrivalent seasonal influenza vaccine; HA, hemagglutinin; RSV, respiratory syncytial virus; WA, Wuhan-Hu-1 (Ancestral).

In Part 1, approximately 120 participants were randomly assigned (2:1:1) to receive either a single dose of mRNA-1345 (50 µg at Day 1), 2 doses of mRNA-1647 (100 µg/dose at Day 1 and Day 57), or 3 doses of mRNA-1647 (100 µg/dose at Day 1, Day 57, and Day 169); data for dose 3 were unavailable at the time of manuscript development. Participants randomly assigned to receive mRNA-1647 were stratified by CMV serostatus in a 2:1 ratio of seronegative to seropositive.

In Part 2, approximately 180 participants were randomly assigned equally to receive either a single dose of mRNA-1010 (50 µg), the active comparator FLUAD (60 µg), or mRNA-1273.222 (50 µg) (both mRNA-1010 and FLUAD were matched to the 2022-2023 Northern Hemisphere influenza strain).

Eligible participants met the age criteria at the time of consent and were in good health based on review of medical history and physical examination. Participants with recent vaccination (COVID-19, influenza) or recent infection with RSV, SARS-CoV-2, or influenza were excluded. Full eligibility criteria are included in the **Supplementary Methods.**

The protocol was approved by a central institutional review board (Advarra, Columbia, MD). The study was conducted in accordance with the protocol, the principles of the International Council for Harmonization Good Clinical Practice guidelines, the Declaration of Helsinki, and national, state, and local laws/regulations. All participants provided written informed consent for participation in the study, including all evaluations and study procedures specified by the protocol.

### 2.2 Study objectives and endpoints

The primary objective was to evaluate the safety and reactogenicity of each study vaccine. The secondary objective was to evaluate the humoral immunogenicity (ie, geometric mean titer [GMT] and geometric mean fold rise [GMFR] of neutralizing antibody [nAb] responses) of the study vaccines at pre-specified time points (**Supplementary Methods**). Given the overarching exploratory nature of this study, multiple exploratory objectives were developed; preliminary antigen-specific T-cell responses following mRNA-1345 or mRNA-1273.222 vaccination were included in this report. In addition, pro-inflammatory and other soluble mediators of inflammation and innate immunity were assessed (**Supplementary Methods**).

### 2.3 Study vaccines and procedures

mRNA-1273.222 (50 µg) is a bivalent COVID-19 vaccine containing mRNAs encoding the cell membrane–associated SARS-CoV-2 spike glycoprotein (Ancestral and omicron BA.4/5 variant)[20]. mRNA-1345 (50 µg) is an RSV vaccine containing a single mRNA encoding membrane-anchored RSV-F glycoprotein, derived from an RSV-A strain, and stabilized in the preF conformation[8]. mRNA-1647 (100 µg) is a CMV vaccine comprising 6 mRNA sequences: 1 mRNA encodes the full-length glycoprotein B (gB), and 5 mRNAs encode the pentameric subunits gH/gL/UL128/UL130/UL131A [21]. mRNA-1010 (50 µg) is a quadrivalent seasonal influenza vaccine encoding membrane-bound hemagglutinin (HA) surface glycoproteins of 4 influenza strains (A/H1N1, A/H3N2, B/Victoria, and B/Yamagata) recommended by the World Health Organization for cell- or recombinant vaccines or the northern hemisphere 2022-23 influenza season [22]. All 4 mRNA vaccines are lipid nanoparticle (LNP)–encapsulated and have similar LNP formulations and amounts of constituents. The difference in each mRNA vaccine is the vaccine antigen and the antigen dose. FLUAD (60 µg) is a licensed, adjuvanted (MF59), inactivated, quadrivalent seasonal influenza vaccine. All study vaccines were administered as an intramuscular injection (0.5 mL). Participants were followed for ≥12 months after the last vaccine dose.

### 2.4 Safety assessments

Safety assessments included solicited local and systemic ARs within 7 days of vaccine administration, unsolicited adverse events (AEs) within 28 days of each vaccine administration, and medically attended adverse events (MAAEs), serious adverse events (SAEs), adverse events of special interest (AESIs), and AEs leading to study withdrawal from Day 1 to the end of the study. Solicited ARs were recorded by patients using eDiaries.

Solicited local ARs included injection site pain, erythema, and swelling/induration; solicited systemic ARs included fever, headache, fatigue, myalgia, arthralgia, nausea-vomiting, axillary swelling or tenderness on the same side as injection, and chills. Solicited ARs were graded as grades 1 through 4; additional information is provided in the **Supplementary Methods**.

### 2.5 Immunogenicity assessments

Blood samples were collected to evaluate the humoral immunogenicity elicited following study vaccination (**Figure 1** and **Supplementary Methods**). Immunogenicity results are presented for the mRNA-1345, mRNA-1273.222, and mRNA-1010 study arms; data analysis is ongoing for the mRNA-1647 study arm and will be reported separately in due course. Immunogenicity assessments were performed using a microneutralization assay (mRNA-1345), a pseudovirus neutralization assay (mRNA-1273.222), and the HA inhibition (HAI) assay (mRNA-1010). Results are presented as GMT or geometric mean concentration (GMC) and GMFR (the geometric mean ratio of post-baseline/baseline titers). T-cell analysis using intracellular cytokine staining was performed using data from a randomized subset of 30 participants in the mRNA-1345 group, divided equally by age cohort, and using data from 57 participants in the mRNA-1273.222 group.

### 2.6 Multiplex cytokine analysis

Serum cytokines, chemokines, and other soluble biomarkers of innate immune activation were analyzed using the Luminex xMAP^®^ and/or Quanterix Simoa^TM^ platform (Rules Based Medicine, Austin, TX). For the Luminex xMAP^®^ analysis, the target cytokine-specific capture antibody was conjugated covalently to a unique set of microspheres. The capture antibody-coupled microspheres were incubated with serum samples for the assay-specific capture antibody on each microsphere to bind to the cytokine of interest. A cocktail of biotinylated detecting reagents was added to the microsphere, followed by a streptavidin-labeled fluorescent “reporter” molecule. The microsphere mixture was analyzed using the Luminex 200™ instrument to determine the concentration of cytokines.

For the Quanterix Simoa^TM^ analysis, the capture antibody conjugated paramagnetic beads were incubated with standards, serum samples, and biotinylated detection antibodies. After washing, the beads were incubated with streptavidin-ß-galactosidase, followed by loading into the Simoa Disc with enzyme substrate, resorufin ß-galactopyranoside. The fluorescence signals were compared with the standard curve, and the quantity of target cytokine was determined for each sample.

### 2.7 Statistical analysis

There was no formal hypothesis testing. Overall, the plan was to enroll 300 participants into the study; this sample size was considered sufficient to provide a descriptive summary of the safety, reactogenicity, and immunogenicity of the study vaccines.

All analyses and data summaries/displays were performed by vaccination arm and for the total population (where applicable) unless otherwise specified; data are presented as categorical variables, frequencies, and percentages. Continuous variables were summarized using descriptive statistics (number of participants, mean, median, standard deviation, minimum, and maximum). All safety analyses were descriptive in nature. For the immunogenicity assessments, seroresponse rate (SRR) from baseline and 2-sided 95% confidence intervals (CIs) were determined using the Clopper-Pearson method at each post-baseline time point. The geometric mean of specific antibody titers with corresponding 95% CIs at each time point and the GMFR of specific antibody titers with corresponding 95% CIs at each post-baseline time point compared with pre-injection baseline at Day 1 were calculated by vaccination arm. For the analysis of mRNA-1010 and FLUAD, the SRR was defined as the proportion of participants with either a pre-vaccination HAI titer <1:10 and a post-vaccination HAI titer ≥1:40 or a pre-vaccination HAI titer ≥1:10 and a minimum 4-fold rise in post-vaccination HAI antibody titer. SRR for humoral immunogenicity (functional and binding antibody titer) was defined as the proportion of participants with a change in the Day 29 (mRNA-1273.22, mRNA-1345) titer of a ≥4-fold rise from baseline. Peak cytokine dynamic was defined as the log_2_ transformed ratio between peak post-baseline cytokine (Day 2) response and baseline (ie, log_2_[post/baseline]). Area under the curve (AUC) of cytokine response over time was calculated based on the trapezoidal rule. The normalized AUC (nAUC) was calculated as the AUC divided by the time range in days. Associations between cytokine response and reactogenicity were quantified via Spearman’s correlation coefficients. Reactogenicity scores were calculated by adding all toxicity grades per AR occurring within 7 days of vaccination administration. The local symptom score was derived by summing all local AR scores; the systemic symptom score was derived by summing all systemic AR scores. Analyses were performed using SAS^®^, version 9.4. Further information on the analysis populations can be found in the **Supplementary Methods.**

## 3 Results

### 3.1 Participants

At the interim analysis of this phase 1 trial (data cutoff February 1, 2023), a total of 308 participants were enrolled and were randomly assigned to receive 1 of the study vaccines. Participants received a single dose of mRNA-1345 (n=61), mRNA-1273.222 (n=60), mRNA-1010 (n=60), or FLUAD (n=59) (**Figure S1**). In the mRNA-1647 group, 62 participants were randomly assigned and received the first dose; 57 and 24 participants received the second and third doses, respectively. Across all groups, 25 participants discontinued the study; no participant discontinued due to safety reasons. Among participants scheduled to receive >1 study vaccine dose (combined mRNA-1647 groups), 7 of 62 (11.3%) discontinued the study vaccine following the first or second injection. The most common reasons for study and vaccine discontinuation were lost to follow-up and participant withdrawal of consent (**Figure S1**).

Across all groups, the mean age was 44.5 (range, 18-73) years; 182 participants were in the group aged 18 to 49 years and 120 participants were in the group aged 50 to 75 years (**Table 1**). Most participants were White (74.8%) and 59.6% were female. Across all groups, more than half of the participants were CMV-positive at baseline. Demographic characteristics between the mRNA-1010 and FLUAD groups were well matched.

**Table 1.**
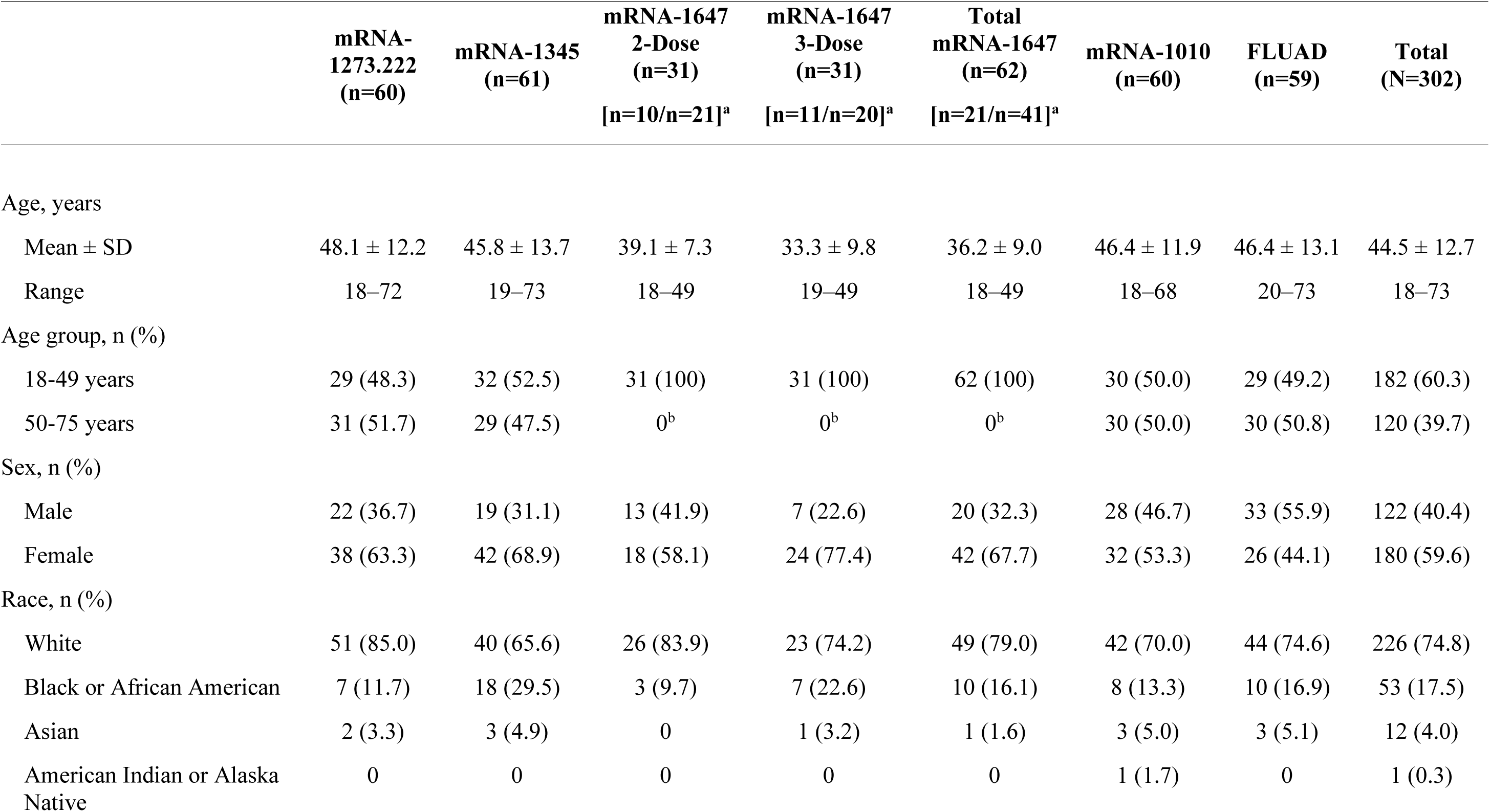
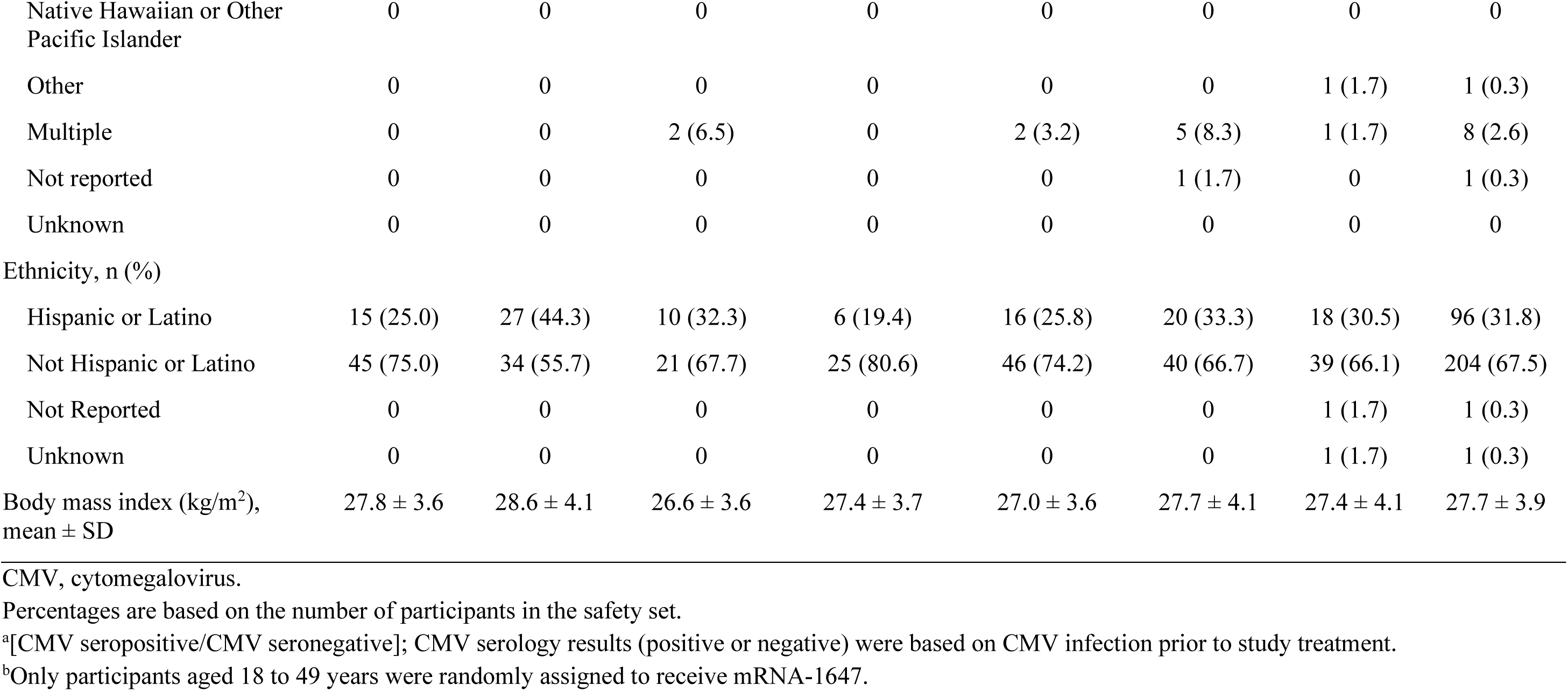
Participant baseline demographics and CMV serostatus (safety set)

### 3.2 Safety

#### 3.2.1 Solicited local adverse reactions

The participant incidence of solicited local ARs ranged from 55.9% to 72.9% across single-dose groups (mRNA-1345, mRNA-1273.222, mRNA-1010, and FLUAD; **Figure 2A**) and was 72.6% and 75.4% after the first and second mRNA-1647 injection respectively (**Figure 2B**). Injection site pain was the most frequently reported solicited local AR for both single-dose (51.7%-72.9%) and mRNA-1647 groups (67.7%, 75.4%). All solicited local ARs were grade 1 or 2 in the mRNA-1345, mRNA-1273.222, and mRNA-1010 groups. In general, local ARs were more common after mRNA-1273 than the other groups but most were Grade 1. Grade 3 solicited local ARs occurred in 1 participant in the FLUAD group and in 4 participants in the mRNA-1647 group (dose 1, n=1; dose 2, n=1; dose 3, n=2). No grade 4 solicited local ARs were reported. The incidence of solicited local ARs was higher among participants aged 18 to 49 years compared with those aged 50 to 75 years in the mRNA-1273.222 (79.3% vs 64.5%) and mRNA-1010 groups (66.7% vs 56.7%) and was lower in the mRNA-1345 (53.1% vs 58.6%) and FLUAD groups (51.7% vs 60.0%) (**Table S1**). In the combined mRNA-1647 groups, the incidence of solicited local ARs was similar after dose 1 and dose 2 (72.6% and 75.4%). The incidence of solicited local ARs was lower among CMV-seropositive than CMV-seronegative participants after both the first (66.7% vs 75.6) and second (66.7% vs 79.5%) doses (**Figure S2** and **Table S2**).

**Figure 2.**
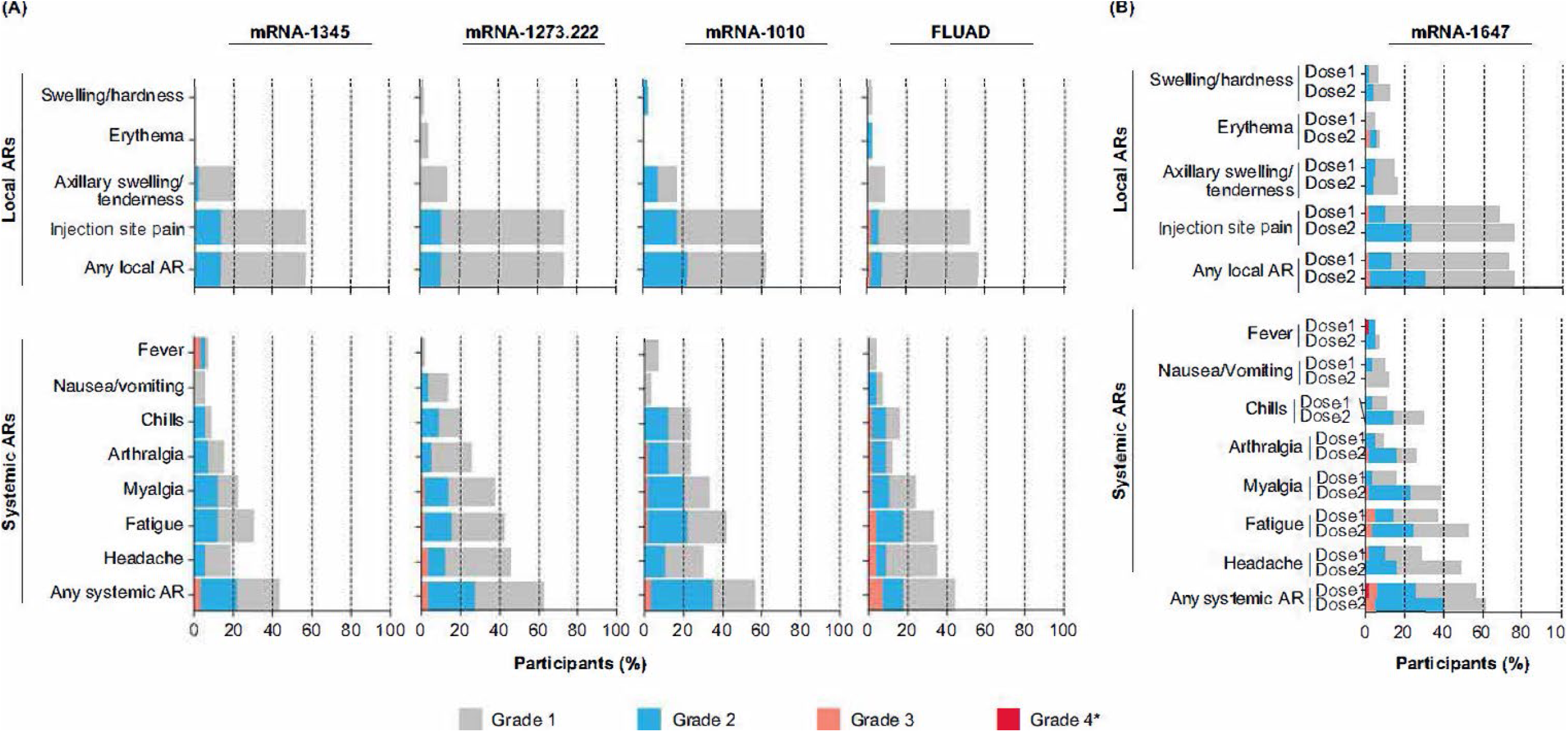
Solicited local and systemic ARs within 7 days of vaccination in (A) the single dose (A) and (B) the 2-and 3-dose groups. Data for the single dose groups (A) are representative of the first injection solicited safety sets for mRNA-1345, mRNA-1273.222, mRNA-1010, and FLUAD. Data for the 2- and 3-dose mRNA-1647 groups (B) were combined and are shown by dose in the first injection solicited safety set (dose 1) and second injection solicited safety set (dose 2) for mRNA-1647. *All grade 4 events were fevers and were the result of self-reported data entry errors and are reported verbatim as recorded in the eDiary. AR, adverse reaction.

Characteristics of solicited local ARs following the first injection are summarized in **Table S3**; median duration ranged from 2 to 3 days for the mRNA groups and was 1 day for the FLUAD group.

#### 3.2.2 Solicited systemic adverse reactions

The participant incidence of solicited systemic ARs ranged from 43.3.% to 62.7% across single -dose groups (mRNA-1345, mRNA-1273.222, mRNA-1010, and FLUAD; **Figure 2A**) and was 56.5 % and 61.4% after the first and second mRNA-1647 injection, respectively (**Figure 2B**). In general, solicited systemic ARs were reported less frequently and were of lower severity in the mRNA-1345 group compared with the other vaccine groups. Across all groups, the most frequently reported solicited systemic ARs were headache, fatigue, and myalgia. The majority of solicited systemic ARs were grade 1 or 2. Grade 3 solicited systemic ARs were reported by 8 participants in the single-dose groups combined, most frequently in the FLUAD group (n=4). In the combined mRNA-1647 groups, 9 participants experienced grade 3 solicited systemic ARs (after dose 1, n=3; after dose 2, n=3; after dose 3, n=3). There were 3 grade 4 solicited systemic ARs that were a result of self-reported data entry errors (mRNA-1345 group, n=2; mRNA-1647 group, n=1 [after dose 1]). The participant incidence of solicited systemic ARs among adults aged 18 to 49 years compared with those aged 50 to 75 years was lower in the mRNA-1345 group (37.5% vs 48.3%), higher in the mRNA-1010 group (60.0% vs 53.3%), and similar in the mRNA-1273.222 (62.1% vs 61.3%) and FLUAD (44.8 vs 43.3%) groups (**Table S1**). In the mRNA-1647 group, the participant incidence of solicited systemic ARs after dose 1 and dose 2 was similar (56.5% and 61.4%); however, the proportion of participants experiencing headache, fatigue, myalgia, arthralgia, and chills was higher after dose 2 than after dose 1 (>10% difference; **Figure S2**).

Characteristics of solicited systemic ARs following the first injection are summarized in **Table S3**; the median duration ranged from 1 to 2 days for the mRNA groups and was 2 days for the FLUAD group.

#### 3.2.3 Unsolicited adverse events

Within 28 days after injection, TEAEs occurred in 13.1% of participants in the mRNA-1345 group, 18.3% in the mRNA-1273.222 group, 8.3% in the mRNA-1010 group, 25.4% in the FLUAD group, and 40.3% in the mRNA-1647 groups combined (**Table S4**). There was a total of four grade ≥3 TEAEs, consisting of 1 event of ischemic stroke in the mRNA-1345 group, 1 event each of COVID-19 pneumonia and osteoarthritis in the FLUAD group, and 1 event of appendicitis in the mRNA-1647 groups combined; none of these events were considered treatment-related.

Treatment-related TEAEs occurred in 1 of 61 participants (1.6%) in the mRNA-1345 group, 4 of 60 (6.7%) in the mRNA-1273.222 group, 0 of 60 (0%) in the mRNA-1010 group, 1 of 59 (1.7%) in the FLUAD group, and 15 of 62 (24.2%) in the mRNA-1647 group (**Table S5**).

There were no AESIs, treatment-related severe TEAEs, or treatment-related SAEs. There were no deaths and no serious, severe, or treatment-related AEs leading to discontinuation from the study or the study vaccine.

### 3.3 Immunogenicity

#### 3.3.1 Humoral immunogenicity

##### 3.3.1.1 mRNA-1273.222

After a single mRNA-1273.222 injection, nAb concentration against all SARS-CoV-2 variants increased from baseline at Day 8, peaked at Day 15, and remained above baseline at Day 29 for both age groups (**Figure 3A-C**). The increase in nAb responses was similar between the younger and older age groups. GMCs were lower for omicron-BA.1 and omicron-BA.4/5 compared with the ancestral variant (**Figure 3A-C**). Antibody concentrations by SARS-CoV-2 serostatus are shown in **Figure S3** and **Supplementary Results**. SRR values are shown in **Table S6** and **Supplementary Results**.

**Figure 3.**
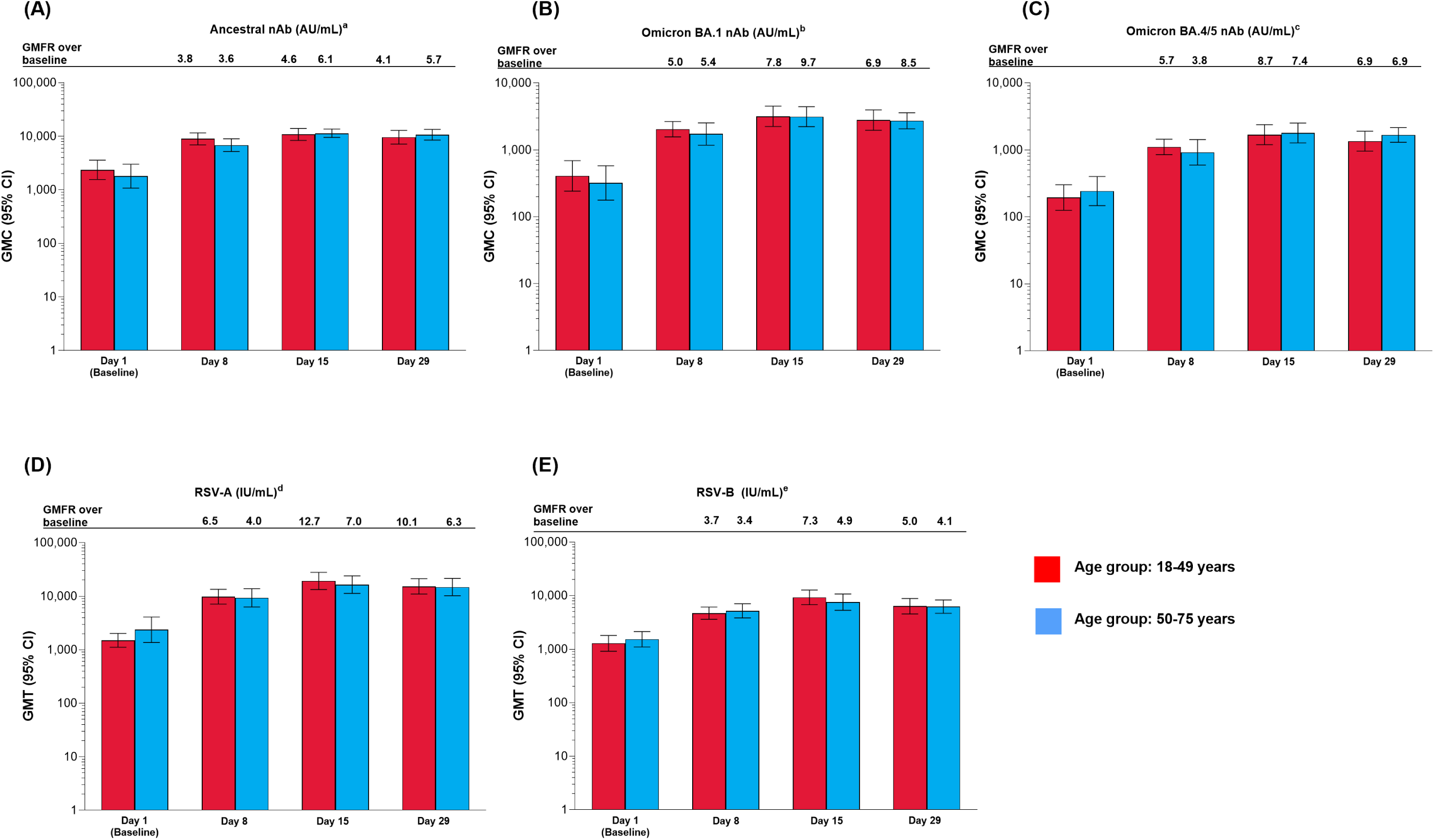
Antibody levels for participants vaccinated with mRNA-1273.222 and mRNA-1345 by age group against A) Ancestral; B) Omicron BA.1; C) Omicron BA.4/5; D) RSV-A (VR-1540); E) RSV-B (per-protocol set). CI, confidence interval; GMFR, geometric mean fold rise, GMT, geometric mean titer; LLOQ, lower limit of quantification; nAb, neutralizing antibody; ULOQ, upper limit of quantification. ^a^LLOQ: 10, ULOQ: 111,433. ^b^LLOQ: 8, ULOQ: 41,984. ^c^LLOQ: 103, ULOQ: 28,571. ^d^LLOQ: 9.94332, ULOQ: 190,682. ^e^LLOQ: 5.5488, ULOQ: 135,524. Antibody values reported as below the LLOQ are replaced by 0.5 × LLOQ. Values greater than the ULOQ are converted to the ULOQ. For baseline (Day 1), GMT with 95% CIs were estimated using a *t*-test. For post-baseline time points, GMTs with 95% CIs were estimated based on a mixed model for repeated measures, with baseline log_10_ titer and day as fixed effects and participant as a random effect. Participants missing SARS-CoV-2 serology sample test results at baseline were excluded from the figure.

##### 3.3.1.2 mRNA-1345

After a single dose of mRNA-1345, neutralizing antibody titers against RSV-A and RSV-B for both age groups were boosted at Day 8, peaked at Day 15, and remained high at Day 29 (**Figure 3D-E**). Peak GMTs were generally higher in younger adults. (**Figure 3D-E** and **Supplementary Results**). SRR values are shown in **Table S7** and **Supplementary Results**.

##### 3.3.1.3 mRNA-1010 and FLUAD

After a single injection of mRNA-1010 or FLUAD, anti-HA antibody levels were boosted at Day 8, peaked at Day 15, and remained elevated at Day 29 for all strains assessed, in both age groups (**Figure 4A-B**). Post-vaccination titers tended to be highest against A-H1N1 compared with the other influenza strains. GMTs and GMFRs were similar following a single dose of mRNA-1010 or FLUAD vaccination and were generally higher in the younger group compared with the older group (**Figure 4A-B**). GMTs and GMFRs were generally higher in the younger group compared with the older group (**Figure 4A-B**). SRRs at Day 29 were similar between the mRNA-1010 and FLUAD groups except for B/Victoria which was lower in the mRNA-1010 group (**Table S8**). A similar trend was observed in both the younger and older cohorts.

**Figure 4.**
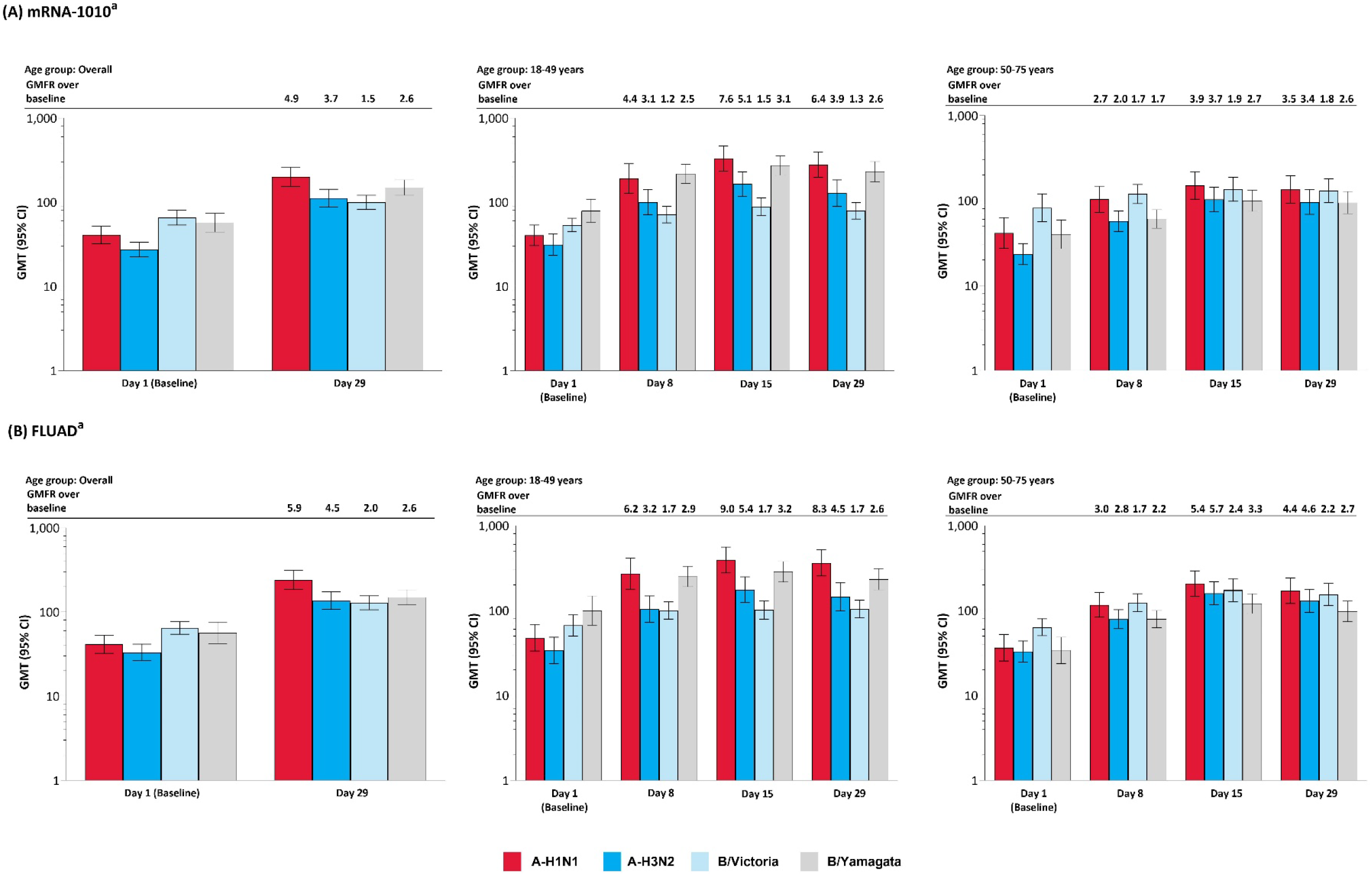
Anti-HA antibody levels by age group in the A) mRNA-1010 group; B) FLUAD group (per-protocol set). A-H1N1, A/Wisconsin/588/2019 (H1N1) -like virus; A-H3N2, A/Darwin/6/2021 (H3N2)-like virus; B/Vict, B/Austria/1359417/2021 (B/Victoria lineage)-like virus; B/Yam, B/Phuket/3073/2013 (B/Yamagata lineage)-like virus; CI, confidence interval; GMFR, geometric mean fold rise; GMT, geometric mean titer; HA, hemagglutinin; LLOQ, lower limit of quantification; ULOQ, upper limit of quantification ^a^Antibody values reported as below the LLOQ are replaced by 0.5 × LLOQ. Values greater than the ULOQ are converted to the ULOQ. For baseline (Day 1), GMTs with 95% CIs were estimated using a *t*-test. For post-baseline time points, GMTs with 95% CIs for mRNA-1010 and FLUAD groups were estimated based on a mixed model of repeated measures, with baseline log_10_ titer, vaccination group, day, and vaccination group by day interaction as fixed effects and participants as a random effect. Participants missing influenza serology sample test results at baseline were excluded from the figure. Influenza A H1N1 antibody (titer): LLOQ: 10, ULOQ: 1280; Influenza A H3N2 antibody (titer): LLOQ: 10, ULOQ: 2560; Influenza B/Victoria-lineage (titer): LLOQ: 10, ULOQ: 640; Influenza B/Yamagata-lineage (titer): LLOQ: 10, ULOQ: 2560).

#### 3.3.2 Cellular immunogenicity

##### 3.3.2.1 mRNA-1273.222

In the combined age cohort, a single dose of mRNA-1273.222 increased preexisting antigen-specific CD4^+^ and CD8^+^ T-cell responses from baseline through Day 29 to the omicron-XBB.1.5 (**Figure 5A**) and ancestral and omicron-BA.4/5 variants (**Figure S4A**). At baseline, CD8^+^ T-cell responses were lower compared with CD4^+^ T-cell responses but showed a greater increase at Day 29 post-vaccination to all strains tested (**Figure 5A** and **Figure S4A**).

**Figure 5.**
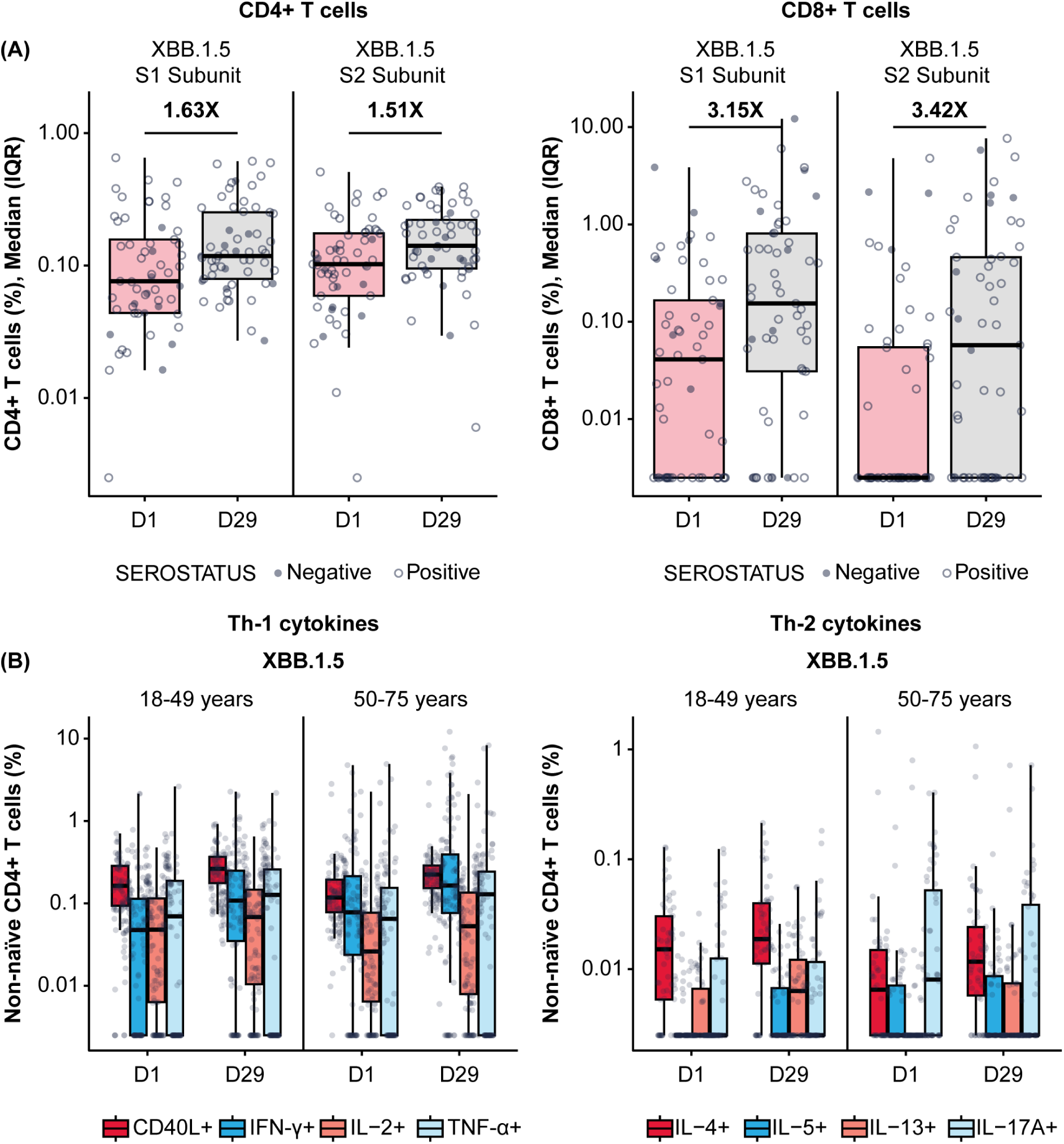
Cellular immunogenicity against omicron-XBB.1.5 following vaccination with mRNA-1273.222. (A) Overall T cell responses (age groups combined); (B) Th-specific T cell responses by age group. ^a^CD4^+^ and CD8^+^ T-cell subsets were gated on CD69^+^ and IFN-γ; assays used were research grade. ^b^Each graph represents the sum of the S1 and S2 peptide pools, which collectively comprise the spike protein. ^c^N=57 participants/time point. ^d^All data was background subtracted from the DMSO control. CD, cluster of differentiation; DMSO, dimethyl sulfoxide; IFN-γ, interferon-γ; IL, interleukin; IQR, interquartile range; LLOQ, lower limit of quantification; S, spike protein subunit; Th1, type 1 helper T cell; Th2, type 2 helper T cell; TNF-α, tumor necrosis factor-α. Th-specific T-cell responses by age group represent the sum of the S1 and S2 peptide pools, which collectively comprise the spike protein. Data are representative of 57 participants per time point. Data were background subtracted from the DMSO control and the assay LLOD was set to 0.005. The solid black data points represent boxplot outliers.

At Day 29 post-vaccination with mRNA-1273.222, CD4^+^ T-cell responses were largely T helper (Th)1–directed (expressing interferon [IFN]-γ, tumor necrosis factor [TNF]-α, interleukin [IL]-2, and CD40L [costimulatory molecule]) in both younger and older adults in response to stimulation with spike peptides from the omicron-XBB.1.5 variant (**Figure 5B**) and ancestral and omicron-BA.4/5 variants (**Figure S4B**). Additionally, a marginal CD4^+^ Th2-directed response (expressing IL-4 and IL-13) was observed through Day 29 in both age cohorts in response to spike peptide stimulation (**Figure S4B**).

##### 3.3.2.2 mRNA-1345

Among mRNA-1345 recipients included in the T-cell analysis (n=30), the percentage of IFN-γ– positive RSV preF-specific CD4^+^ or CD8^+^ T-cells was low or undetectable prior to vaccination (Day 1) (**Figure S5**). The percentage of IFN-γ–positive RSV preF-specific CD4^+^ or CD8^+^ T cells increased from baseline at Day 8 post-vaccination, peaked at Day 15, and remained above baseline through Day 91 (**Figure S5A**). Peak CD4^+^ T-cell responses were numerically lower in older adults (50-75 years) than younger adults (18-49 years), although relative increases were greater in the former age group (**Figure S5B**). RSV-specific CD4^+^ T cells were enriched for the Th1 (expressing IFN-γ, IL-2, TNF-α, and CD40L) compared with Th2 phenotype, with similar frequencies observed for IFN-γ and TNF-α given the assay response variability (**Figure 6A-B**). More than 10-fold higher frequency of Th1 compared with Th2 cytokines was elicited by mRNA-1345 vaccination; the Th2 cytokines were predominantly low-frequency IL-4 responses, whereas IL-5 and IL-13 were either near the lower detection limit or non-detectable for most participants (**Figure 6A-B**). No discernible differences in the cytokine profiles at baseline or after vaccination were observed between the 2 age cohorts (**Figure 6A-B**). Cytokines associated with CD8^+^ T-cell responses consisted mainly of IFN-γ, TNF-α, and IL-2, with the highest frequency observed for IFN-γ. CD8^+^-mediated T-cell responses were generally higher in older adults(**Figure 7**).

**Figure 6.**
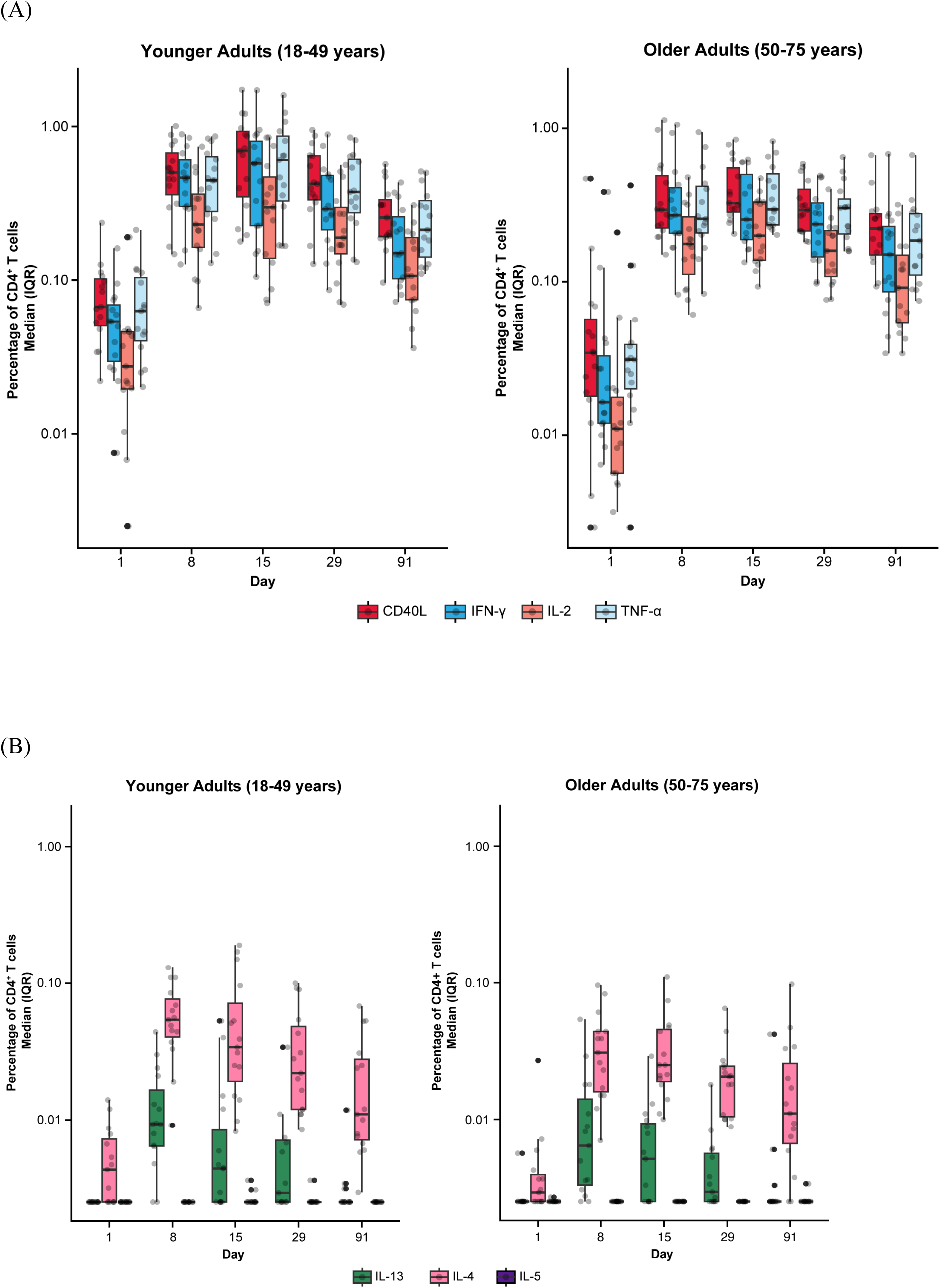
RSV preF-specific CD4^+^ T cells among participants in the mRNA-1345 group by age cohort (A) Th1 response and (B) Th2 response. CD, cluster of differentiation; IFN-γ, interferon-γ; IL, interleukin; ICS, intracellular cytokine staining; IQR, interquartile range; LLOD, lower limit of detection; preF; prefusion; RSV, respiratory syncytial virus; Th1, type 1 helper T cell; Th2, type 2 helper T cell; TNF-α, tumor necrosis factor-α. LLOD for the ICS assay was 0.005%. CD4^+^ T-cell subsets were gated on CD69^+^ and split into the following groups: Th1: IFN-γ, TNF-α, IL-2; Th2: IL-4, IL-5, IL-13; and costimulatory T cells: CD40L; assays used were research grade. The solid black data points represent boxplot outliers.

**Figure 7.**
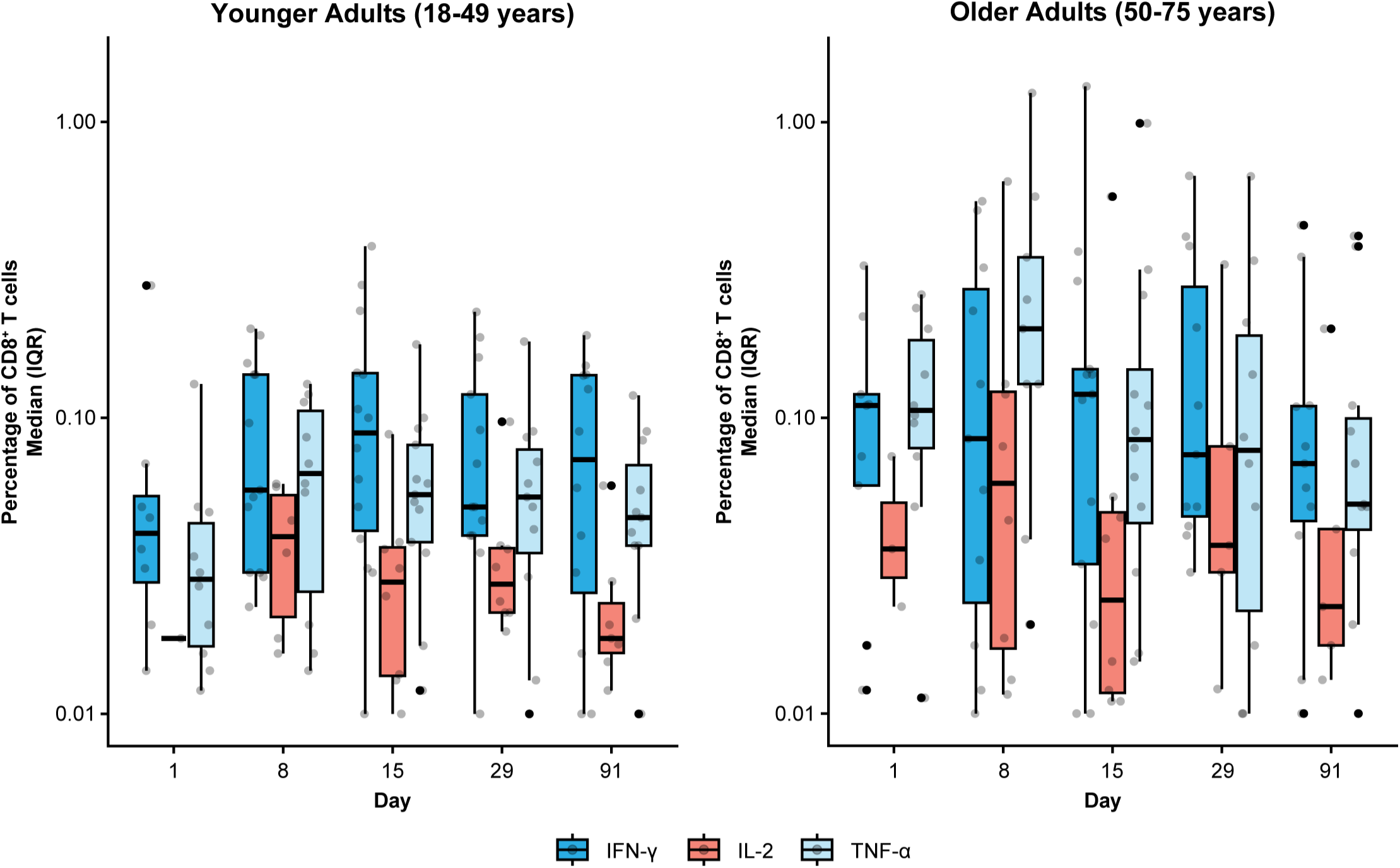
RSV preF-specific CD8^+^ T cells for the mRNA-1345 group by age cohort. CD, cluster of differentiation; IFN-γ, interferon-γ; IL, interleukin; IQR, interquartile range; preF; prefusion; RSV, respiratory syncytial virus; TNF-α, tumor necrosis factor-α. CD8^+^ T cell subsets were gated on CD69^+^: IFN-γ, TNF-α, IL-2; assays used were research grade. The solid black data points represent boxplot outliers.

### 3.4 Biomarkers of innate immune activation

The chemokine/cytokine response analysis showed a distinct signature with mRNA vaccines compared with FLUAD (**Figure 8**). Of the 63 cytokines detected, IFN-γ, IL-6, IL-2Ra, CXCL9, IP-10, MCP-2, and MIP-1β were elevated on Day 2 compared with baseline (Day 1) (**Figure 8**). CXCL9, IP-10, IL-2Ra, and MCP-2 remained elevated on Day 4; the levels returned to baseline on Day 8 (**Figure 8**). Increased IFN-γ was observed for all vaccine groups, with levels peaking on Day 2 after vaccination and showing a higher magnitude increase with mRNA vaccines (**Figure 8**). IL-6 responses in mRNA vaccine groups were variable, with low magnitude increases observed in some participants at 6 to 12 hours/day after vaccination (**Figure 8**). Across the mRNA vaccine groups, mRNA-1345 was associated with the lowest magnitude cytokine response, which was similar to FLUAD (**Figure 8**). Responses were similar between older and younger adults for all vaccine groups (**Figure 8**). Additional biomarkers that showed increases from baseline on Days 2 to 4 were C-reactive protein (CRP), serum amyloid P (SAP), and β-2 macroglobulin (β2M); conversely, a number of biomarkers showed decreases from baseline (**Figure S6A**). No evidence of response following vaccination was observed for TNF-α, TNF-β, IFN-α, IFN-β, IL-12p70, IL-12p40, and IL-1β) (**Figure S6B**).

**Figure 8.**
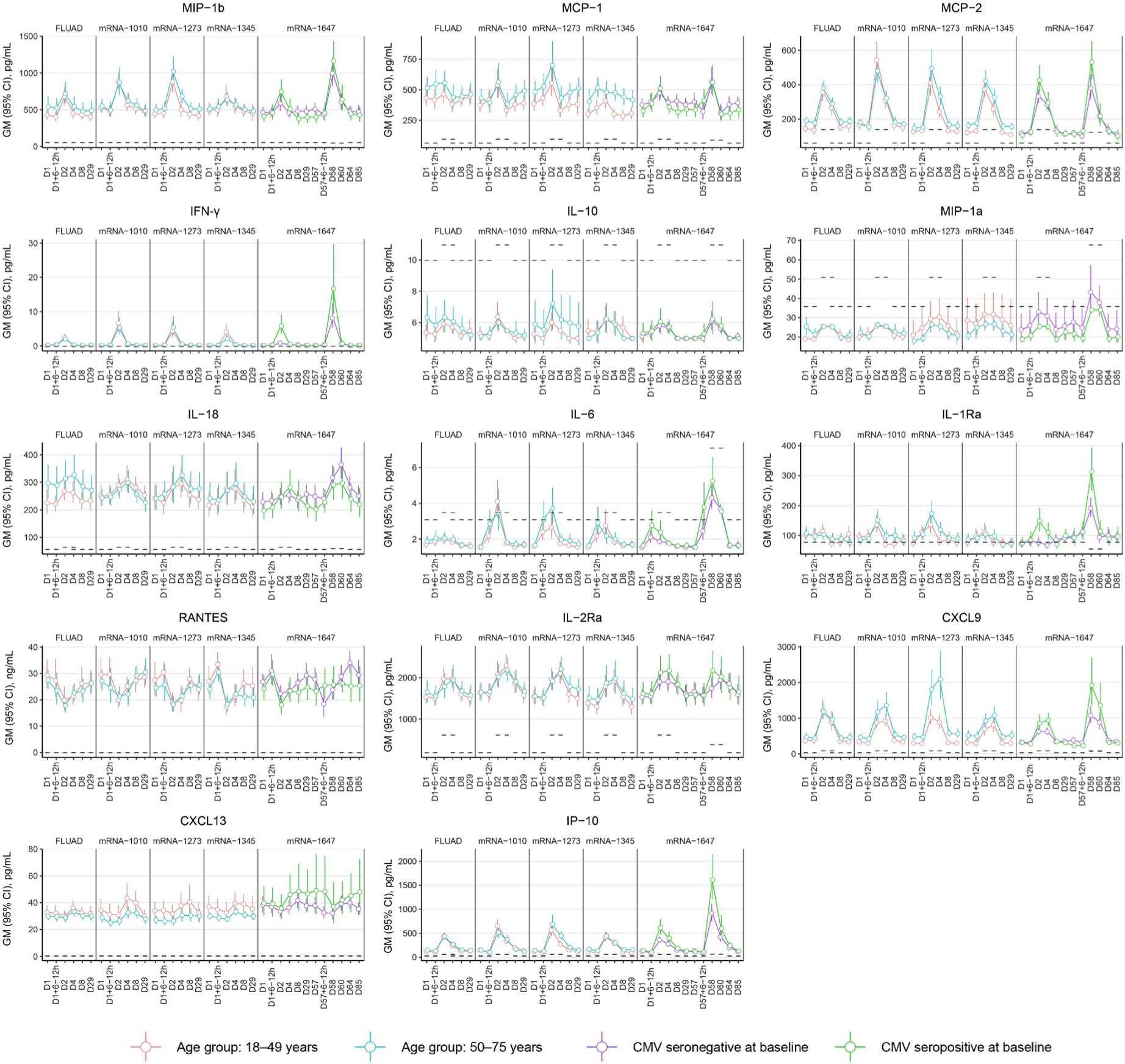
Cytokine response from peripheral blood following vaccination with FLUAD and mRNA vaccines. Geometric mean (95% CI) of cytokine levels after vaccination by the study visit, age group, and CMV status (mRNA-1647 group). Dotted lines represent the LLOQ for each analyte. For some time points, the LLOQ is either decreased or increased, since not all samples could be assessed prior to the establishment of a revised LLOQ for samples run at a later time point. CI, confidence interval; CMV, cytomegalovirus; CRP, C-reactive protein; CXCL9, CXC chemokine ligand 9; CXCL13, CXC chemokine ligand 13; IFN-γ, interferon-γ; IL, interleukin; IL-2Rα, interleukin-2 receptor α; IP-10, interferon γ-induced protein 10 kDa; LLOQ, lower limit of quantification; MCP, monocyte chemotactic protein; MIP-1α, macrophage inflammatory protein−1 α; MIP-1β, macrophage inflammatory protein−1 β; MCP-1, monocyte chemotactic protein 1; MCP-2, monocyte chemotactic protein 2; RANTES, T-cell−specific protein RANTES.

In the mRNA vaccine groups, elevated serum levels of IL-1Ra and MCP-1/MCP-2 on Day 2 correlated positively with the presence of systemic reactions (correlation coefficient range, 0.15-0.27) (**Figure 9A**). In the FLUAD group, a higher correlation magnitude was observed between individual systemic reactogenicity rating and serum peak levels of alpha-1 antitrypsin, complement C3, Vitamin D binding protein, CCL11 (eotaxin-1), haptoglobin, and IL-12p40 (correlation coefficient range, 0.32-0.37) (**Figure 9B**). Different correlation patterns with reactogenicity scores were identified in both mRNA and FLUAD groups when using an nAUC measure to incorporate all longitudinal serum cytokine results (**Figure S7A-B**).

**Figure 9.**
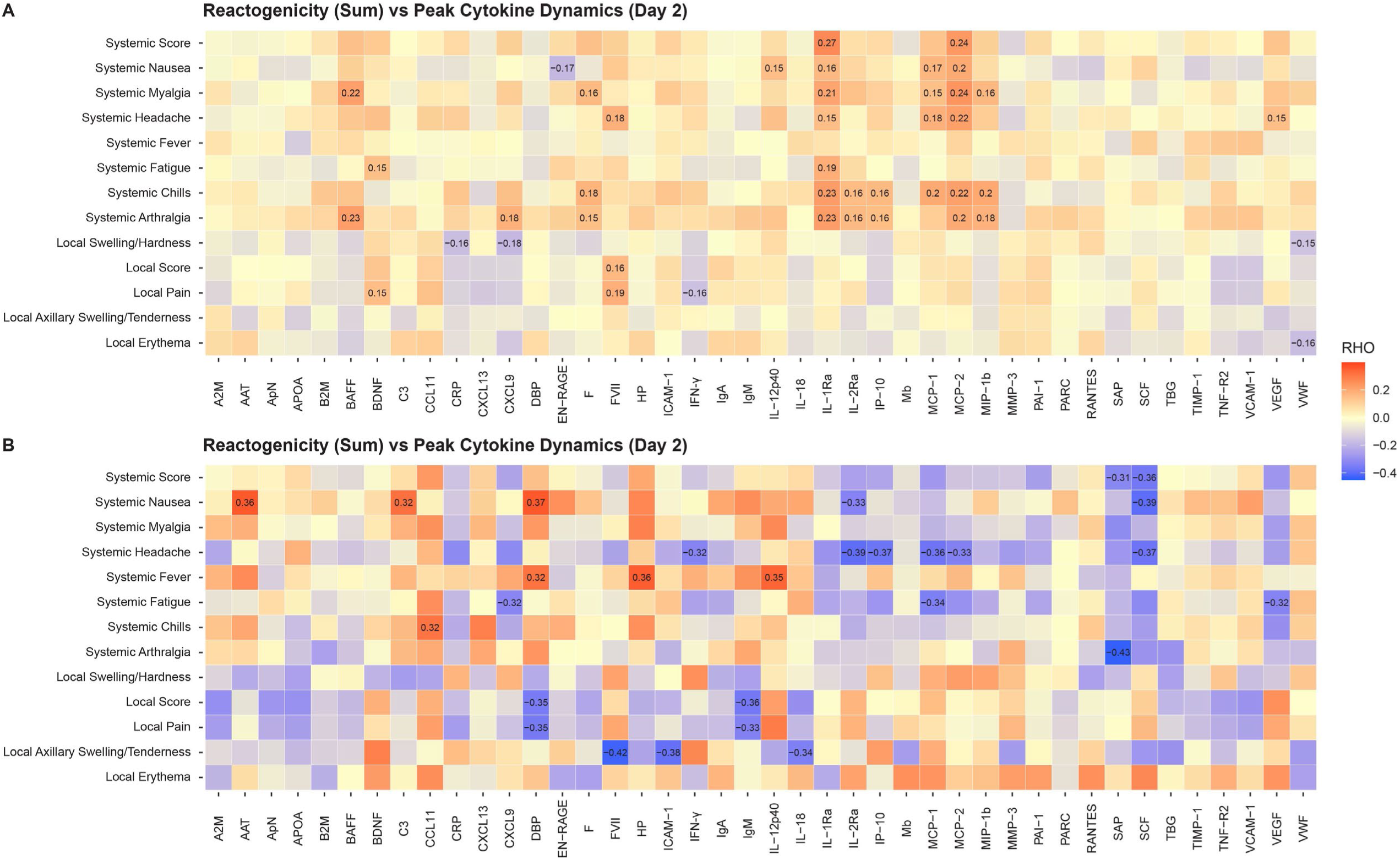
Correlation of peak serum biomarker responses and reactogenicity, (A) mRNA groups, (B) FLUAD group. Correlation of peak serum biomarker responses (Day 2) and peak reactogenicity (highest reported grade) and reactogenicity sum (sum of all grades across symptoms). For all mRNA vaccine groups combined and the adjuvanted quadrivalent influenza vaccine (FLUAD). Statistically significant values are labeled with the correlation coefficient. A2M, alpha-2-macroglobulin; AAT, alpha-1-antitrypsin; ApN, adiponectin; APOA, apolipoprotein(a); B2M, beta-2-microglobulin; BAFF, B-cell activating factor; BDNF, brain-derived neurotrophic factor; C3, complement C3; CCL11, C-C motif chemokine 11; CRP, C-reactive protein; CXCL9, CXC chemokine ligand 9; CXCL13, CXC chemokine ligand 13; DBP, vitamin D–binding protein; EN-RAGE, extracellular newly identified receptor for advanced glycation end products binding protein; F, ferritin; FVII, factor VII; HP, haptoglobin; ICAM-1, intercellular adhesion molecule 1; IFN-γ, interferon-γ; IgA, immunoglobulin A; IgM, immunoglobulin M; IL-12p40, interleukin-12 subunit p40; IL-18, interleukin-18; IL-1Ra, interleukin-1 receptor antagonist; IL-2Ra, interleukin-2 receptor α; IP-10, interferon gamma-induced protein 10; Mb, myoglobin; MCP-1, monocyte chemotactic protein 1; MCP-2, monocyte chemotactic protein 2; MIP-1b, macrophage inflammatory protein-1 beta; MMP-3, matrix metalloproteinase-3; PAI-1, plasminogen activator inhibitor 1; PARC, pulmonary and activation-regulated chemokine; RANTES, T-cell−specific protein RANTES; SAP, serum amyloid P-component; SCF, stem cell factor; TBG, thyroxine-binding globulin; TIMP-1, tissue inhibitor of metalloproteinases 1; TNF-R2, tumor necrosis factor-receptor 2; VCAM-1, vascular cell adhesion molecule-1; VEGF, vascular endothelial growth factor; VWF, von Willebrand factor.

## 4 Discussion

Here, we report interim findings from an ongoing phase 1 clinical study designed to comprehensively characterize mRNA vaccine innate and adaptive immunity by comparatively studying several mRNA vaccines, including SARS-CoV-2 (bivalent mRNA-1273.222), RSV (mRNA-1345), influenza (mRNA-1010), and CMV (mRNA-1647) to identify both shared and unique clinical and immunologic features of mRNA vaccines. This is the first direct, randomized comparison of mRNA-based vaccines that use similar LNP formulations and chemistry, manufacturing and control processes but encode different antigens.

Across vaccine study arms, safety and humoral immunogenicity findings were consistent with previously reported data for these vaccines [8,20,22–26]. Safety findings demonstrate that vaccine reactogenicity was acceptable across vaccine groups. Most solicited local and systemic ARs within 7 days were mild or moderate in severity (grade 1 or 2). These findings are mostly in agreement with larger studies that have reported the safety of mRNA-1273.222, mRNA-1345, mRNA-1010, and mRNA-1647 in adults [8,20,22–26]. However, mRNA-1273 reactogenicity was higher in earlier studies of the primary series when seropositivity was low [3]. Local solicited ARs (Grade 1) were more commonly reported among recipients of mRNA-1273, and systemic symptoms were reported less frequently and of a lower severity among recipients of the mRNA-1345 vaccine compared with the other vaccine groups. Similarly, the majority of systemic reactions reported by older adults (≥60 years) in the pivotal phase 2-3 trial of mRNA-1345 were mild to moderate in severity [8]. The incidence of solicited ARs was similar in the mRNA-1010 and FLUAD groups. For the mRNA-1647 groups, any solicited AR up to 7 days after injection was similar following doses 1 and dose 2; solicited systemic ARs were reported more frequently after dose 2, consistent with observations for the mRNA-1273 primary series [3]. Across all mRNA vaccine groups, there were no AESIs, treatment-related SAEs, or deaths.

Consistent with prior studies, all vaccines assessed elicited strong functional antibody responses [20,22,23,25,26]. At Day 29, antibody titers against vaccine-specific antigens were increased versus baseline for all mRNA vaccines. Following single-dose mRNA-1273.222 vaccination, Day 29 GMFRs demonstrated that nAb titers were significantly boosted relative to baseline against both the ancestral and omicron variants, as previously reported [20], with peak GMTs higher against ancestral compared with the omicron-lineage variants; boosting as measured by GMFR, was similar across strains. In line with results from the RSV mRNA-1345 phase 1 study, vaccination significantly boosted RSV-A and RSV-B nAbs, with a higher GMT observed against RSV-A, which may be due to assay differences [25–28]. Vaccination with mRNA-1010 resulted in increased HA titers at Day 29, which were similar to or slightly lower than the enhanced seasonal influenza vaccine, FLUAD, particularly for B/Victoria, which is in line with the results of the phase 1/2 study (NCT04956575) of mRNA-1010 [22]; however, preliminary findings from an ongoing phase 3 study assessing the safety and immunogenicity of an updated formulation of mRNA-1010 (NCT05827978) demonstrated improved immunogenicity against B strains [29,30].

To our knowledge this is the first publication evaluating T-cell responses following the vaccination with SARS-CoV-2-targeting mRNA-1273.222 or RSV-targeting mRNA-1345. Here we showed that both SARS-CoV-2 mRNA-1273.222 and RSV mRNA-1345 increased antigen-specific T cells with a strongly biased Th1 antiviral response, with predominant expression of IFN-γ, TNF-α, and IL-2 along with the B-cell co-stimulatory molecule CD40L (CD154). Vaccination with mRNA-1273.222 increased CD4^+^ and CD8^+^ T-cell responses from baseline through Day 29 against both ancestral and omicron SARS-CoV-2 variants. Among mRNA-1345 recipients, RSV preF-specific CD4^+^ and CD8^+^ T-cells were low or undetectable before mRNA-1345 vaccination and were strongly boosted by Day 8 and remained above baseline through Day 91 after vaccination. This finding is encouraging, as a preclinical study found evidence supporting a key role for primed CD4^+^ and CD8^+^ T-cells in controlling RSV infection [31]. With mRNA-1273.222 vaccination, there was a marginal increase in T cells expressing the Th2 cytokines IL-4 and IL-13. Whereas CD4^+^ T cells expressing the Th2 cytokines IL-5 and IL-13 were either not detected or detected at a very low frequency after mRNA-1345 vaccination and IL-4 was detected at a low frequency. Together, these results demonstrate the ability of mRNA vaccination to boost antigen-specific CD4^+^ and CD8^+^ T-cell responses.

Cytokines and chemokines are crucial for guiding the immune response against infection and following vaccination [32]. For vaccines to effectively prime the immune system to elicit an effective adaptive immune response, it is likely important to generate an initial antiviral response, which is often driven by cytokines such as IFN-γ [33]. In this study, IFN-γ and related cytokines (CXCL9, MCP-1, MCP-2) [34,35] increased the day after vaccination for all vaccine groups, but the increase was more pronounced in individuals who received mRNA vaccines. These findings are in line with prior studies with mRNA vaccines [19]. In a study investigating immune cell dynamics following vaccination, 2-dose SARS-CoV-2 mRNA vaccination led to an increase in IFN-γ levels and subsequent activation of IFN-γ–inducible chemotaxis as initial triggers of the cellular processes associated with nAb responses and systemic AEs [36]. However, the specific cell types involved and the regulation of chemokine receptor expression were different for nAb titers and systemic AEs [36]. Notably, the vaccine-induced chemokine receptor (CCR2 or CXCR3) upregulation on natural killer/monocyte cell subsets enhances the ability of these cells to migrate to inflammatory sites and correlates with high nAb titers [36]. Conversely, dendritic cell subsets exhibiting constitutive expression of these chemokine receptors were linked to systemic AEs [36]. In the current study, a low-moderate, positive correlation was observed between peak serum levels of IL-1Ra and MCP-1/MCP-2 and the majority of systemic and local reactogenicity scores in the mRNA vaccine groups at Day 2. These findings suggest that the reactogenicity following vaccination with mRNA vaccines (as well as FLUAD) may occur due to increased cytokine production, albeit at very low levels (often just above the level of detection), leading to an effective immune response [32,37]. Notably, the unique cytokine profile observed with mRNA vaccines relative to FLUAD point to the LNP formulation/adjuvant as the differentiator between mRNA and FLUAD vaccine platforms. It is also important to note that the potentially beneficial effect of reactogenicity (as a clinical indicator of the innate immune response to vaccination) should be balanced or minimized to achieve a clinically effective dose level [37]; as noted previously, most solicited local and systemic ARs within 7 days of mRNA vaccination were mild or moderate in severity.

Aging is known to significantly impact vaccine effectiveness and immunogenicity [38], and the effect of aging on mRNA vaccine response was a key component of the study design. In this study, some differences in safety and immunogenicity were observed with younger versus older participants. The incidence of any solicited AR up to 7 days after injection was slightly higher among younger than older participants in the mRNA-1273.222 and mRNA-1010 groups and was slightly lower for participants in the mRNA-1345 and FLUAD groups. The findings for mRNA-1345 vaccination differ from a previous phase 1 study that reported a higher incidence of solicited local and systemic ARs in younger versus older adults; it is possible this was due to different definitions of older adults in the 2 studies (phase 1, 65-79 years; present study, 50-75 years) [25] or the subjectivity associated with self-reported symptoms. With RSV mRNA-1345 vaccination, increases in GMTs from baseline to Day 29 were observed against both strains in younger and older adults, with 6- to 10-fold increases for RSV-A and 4- to 5-fold increases for RSV-B, consistent with the findings of the phase 1 study.[25,26] For the influenza vaccine mRNA-1010, similar trends were observed in the older and younger participants, with the older participants having slightly lower titers overall, which has been reported previously in older adults following influenza vaccination [22,39]. In contrast, antigen-specific T-cell responses, including CD8^+^ T cells, were similar or very modestly lower in older versus younger adults for mRNA-1273.222 and RSV mRNA-1345. Furthermore, the cytokine response was similar between older and younger participants for all vaccine groups, with more pronounced IFN-γ upregulation observed with mRNA vaccination, and a potential advantage of mRNA vaccines compared with other vaccine platforms.

This study had several strengths and limitations. This study was limited by the small number of participants in each vaccination group; a small number of participants were included due to the study’s complexities. The study involved frequent participant visits and blood draws of both serum and polymorphonuclear blood cells to support the exploratory nature of the research and advance understanding of the mRNA/LNP vaccine platform. However, the quality and frequency of the sample collections was leveraged to generate additional clinical data on antigen-specific T-cell responses, which we report here for mRNA-1273.222 and mRNA-1345 vaccination. These data are important as they may provide insight into the potential adaptive response and antiviral activity of the immune responses to mRNA vaccines [40].

In conclusion, this interim analysis of the safety and immunogenicity of 4 mRNA vaccines, mRNA-1273.222 (SARS-CoV-2), mRNA-1345 (RSV), mRNA-1010 (influenza), and mRNA-1647 (CMV), raised no safety concerns, with no vaccine-related SAEs or AESIs, and vaccine reactogenicity was acceptable for the 4 mRNA vaccines studied. Immunogenicity findings confirmed that each vaccine was immunogenic against their respective antigens in adults. Antigen-specific T cells were increased with mRNA-1273.222 and mRNA-1345 vaccination, and antigen-specific Th1 responses were enriched. mRNA-1010 was able to elicit similar responses to FLUAD, an adjuvanted influenza vaccine. Further, we demonstrate that mRNA vaccination results in a consistent inflammatory/innate immune profile, similar to the adjuvanted comparator vaccine, which peaks the day after vaccination, commensurate with the median onset of systemic symptoms, with slightly lower peak cytokine levels in the mRNA-1345 group, which reported fewer overall systemic symptoms compared with the other mRNA vaccination groups. A positive correlation between peak cytokine levels (IL-1Ra and MCP-1/MCP-2) and the presence of systemic reactogenicity reactions following mRNA vaccination was also demonstrated, indicative of an increased immune response [37].

Together, these results suggest that the observed similarity in reactogenicity and immunogenicity of mRNA vaccines may be related to shared features of the mRNA platform. Evaluated vaccines demonstrated enhanced antigen-specific CD4+ and CD8+ T-cell responses with a correlation between peak cytokine levels and reactogenicity scores observed on day 2. Further systems biology analyses of this study are ongoing or planned, including using high-dimensional laboratory workflows and assays to further assess and compare the contribution of the innate and adaptive immune responses to each of these vaccines as well as integrated analyses. Overall, this study serves as a basis to further our understanding of mRNA-based vaccination in parallel with the continued development of mRNA vaccines against infectious diseases, which will lead to further improvements in vaccine tolerability, effectiveness, and durability.

## Data Availability

As the trial is ongoing, access to patient-level data presented in the article and supporting clinical documents by qualified external researchers who provide methodologically sound scientific proposals may be available upon reasonable request for products or indications that have been approved by regulators in the relevant markets and subject to review from 24 months after study completion. Such requests can be made to Moderna, Inc., 325 Binney Street, Cambridge, MA 02142. A materials transfer and/or data access agreement with the sponsor will be required for accessing shared data. All other relevant data are presented in the paper. The protocol is available online at ClinicalTrials.gov: NCT05397223

## Author Contributions

CF: Writing - Review & Editing. NS-C: Writing - Review & Editing. DM: Writing - Review & Editing. WZ: Writing - Review & Editing. YS: Writing - Review & Editing. PR: Formal Analysis, Writing - Review & Editing. BG: Writing - Review & Editing. AA: Formal Analysis, Writing - Review & Editing. AD: Conceptualization, Formal Analysis, Writing - Review & Editing. RP: Conceptualization, Formal Analysis, Writing - Review & Editing. All the authors met the authorship criteria and approved the publication.

## Acknowledgments

The authors would like to acknowledge the trial participants, trial investigators, the clinical trial site, and Moderna and Parexel study personnel for their contributions to the study. Medical writing and editorial assistance were provided by Syed Abdul Haseeb, MS, of MEDiSTRAVA in accordance with Good Publication Practice (GPP 2022) guidelines, funded by Moderna, Inc., and under the direction of the authors.

