## Supplement for "Shared Clinical and Immunologic Features of mRNA Vaccines: Preliminary Results from a Comparative Clinical Study"

**Supplementary Information**

**Supplementary Methods**

**Study inclusion criteria**

Each participant must meet all of the following criteria to be enrolled in this study:

1. Adults 18 to 75 years of age at the time of consent (screening visit) who, in the opinion of the investigator, were in good health based on review of medical history and physical examination performed at screening. For mRNA-1647 study arms, adults 18 to < 50 years of age at the time of consent (screening visit) who, in the opinion of the investigator, were in good health based on review of medical history and physical examination performed at screening.
2. Investigator assessment that participant understands and was willing and physically able to comply with protocol-mandated follow-up, including all procedures.
3. Participant had provided written informed consent for participation in this study, including all evaluations and procedures as specified in the protocol.
4. Body mass index of 18 kg/m^2^ to 35 kg/m^2^ (inclusive) at the screening visit.
5. Female participants of nonchildbearing potential were enrolled in the study. Nonchildbearing potential was defined as bilateral tubal ligation >1 year prior to screening, bilateral oophorectomy, hysterectomy, or menopause. A follicle-stimulating hormone level was measured, as necessary, and at the discretion of the investigator, to confirm postmenopausal status.
6. Female participants of childbearing potential were enrolled in the study if the participant fulfilled all the following criteria:

- Had a negative pregnancy test at the screening visit and on the day of vaccination.
- Had practiced adequate contraception or had abstained from all activities that could result in pregnancy for at least 28 days prior to Day 1. Adequate female contraception was defined as consistent and there is correct use of an US Food and Drug Administration–approved contraceptive method in accordance with the product label.
- Had agreed to continue adequate contraception through 3 months following the last study vaccine administration.
- Was not currently breastfeeding.

**Study exclusion criteria**

Participants meeting any of the following criteria will be excluded from the study:

1. Participant have had close contact to someone with confirmed SARS-CoV-2, RSV, or influenza infection in the past 14 days prior to the screening visit.
2. Clinical screening laboratory values (total white blood cell count, hemoglobin, platelets, alanine aminotransferase, aspartate aminotransferase, creatinine, alkaline phosphatase, and total bilirubin) that were grade 2 or above.
3. Participant was acutely ill or febrile (temperature ≥38.0°C/100.4°F) 72 hours prior to or at the screening visit or Day 1. Participants meeting this criterion were rescheduled within the 28-day screening window and retained their initially assigned participant number.
4. History of a diagnosis or condition that, in the judgment of the investigator, was clinically unstable or may affect participant safety, assessment of safety endpoints, assessment of immune response, or adherence to study procedures. Clinically unstable was defined as a diagnosis or condition requiring significant changes in management or medication within the 2 months prior to screening and includes ongoing workup of an undiagnosed illness that could lead to a new diagnosis or condition.
5. History of myocarditis, pericarditis, or myopericarditis.
6. Reported history of congenital or acquired immunodeficiency, to include human immunodeficiency virus, an immunosuppressive condition, or an immune-mediated disease requiring immunosuppressive therapy.
7. Dermatologic conditions that could affect local solicited AR assessments (eg, tattoos, psoriasis patches affecting skin over the deltoid areas).
8. Reported history of anaphylaxis or severe hypersensitivity reaction after receipt of an mRNA vaccine or any components of an mRNA vaccine.
9. Severe allergic reaction to any component of the FLUAD vaccine (active comparator), including egg protein, or after a previous dose of any influenza vaccine.
10. Reported history of bleeding disorder that was considered a contraindication to intramuscular injection or phlebotomy.
11. Any medical, psychiatric, or occupational condition, including reported history of alcohol or substance use, that in the opinion of the investigator, might pose additional risk due to participation in the study or could interfere with the interpretation of study results.
12. Participant had received systemic immunosuppressants or immune-modifying drugs for >14 days in total within 6 months prior to screening (to include any systemic corticosteroids) or was anticipating the need for immunosuppressive treatment at any time during participation in the study.
13. Participant planned to receive any licensed or authorized vaccine, including COVID-19 or influenza vaccines, within 28 days before or after any study injection.
14. Participant had received a northern hemisphere 2021-2022 or 2022-2023 seasonal influenza vaccine or any other influenza vaccine, or an experimental RSV or CMV vaccine, within 6 months prior to study Day 1.
15. Participant had received an authorized or approved COVID-19 vaccine within 4 months prior to Day 1, and/or has not completed a primary vaccination series for COVID-19.
16. Participant had a laboratory-confirmed infection with influenza or RSV within 6 months prior to Day 1, or SARS-CoV-2 within 4 months of Day 1. A SARS-CoV-2 infection confirmed with self-administered, authorized, or approved rapid antigen test was acceptable.
17. Participant had received systemic immunoglobulins or blood products within 3 months prior to the screening visit or plans to receive the treatment during the study.
18. Diagnosis of malignancy within the previous 10 years (excluding nonmelanoma skin cancer).
19. Participant had donated ≥450 mL of blood products within 28 days prior to the screening visit or planned to donate blood products during the study.
20. Participated in an interventional clinical study within 28 days prior to the screening visit based on the medical history interview or plans to do so while participating in this study.
21. Participant was an immediate family member or household member of study personnel, study site staff, or Sponsor personnel.

**Time points for secondary and exploratory endpoints**

Pre-specified time points for measuring humoral immunogenicity (ie, GMT and GMFR of neutralizing antibody responses) were Days 1, 8, 15, and 29 for participants in the single-dose groups (mRNA-1273.222, mRNA-1010, FLUAD, and mRNA-1345 groups), and Days 1, 8, 15, 29, 57, 64, 71, and 85 for participants in the mRNA-1647 group. Prespecified time points for the immunogenicity assessments were Days 1, 8, 15, 29, and 91 for participants in the mRNA-1345 group and Days 1 and 29 in the mRNA-1273.222 group.

**Safety assessments**

Grading for solicited ARs was based on grading scales modified from the Toxicity Grading Scale for Healthy Adult and Adolescent Volunteers Enrolled in Preventive Vaccine Trials.

| **Reaction** | **Grade 1** | **Grade 2** | **Grade 3** | **Grade 4** |
| --- | --- | --- | --- | --- |
| Injection site pain | Does not interfere with activity | Interferes with activity | Prevents daily activity | Requires emergency department visit or hospitalization |
| Injection site erythema (redness) | 25-50 mm/  2.5-5 cm | 51-100 mm/  5.1-10 cm | >100 mm/  >10 cm | Necrosis or exfoliative dermatitis |
| Injection site swelling/  induration (hardness) | 25-50 mm/  2.5-5 cm | 51-100 mm/  5.1-10 cm | >100 mm/  >10 cm | Necrosis |
| Axillary (underarm) swelling or tenderness ipsilateral to the side of injection | No interference with activity | Some interference with activity | Prevents daily activity | Emergency department  visit or hospitalization |
| Headache | No interference with activity | Some interference with activity | Prevents daily activity | Requires emergency department  visit or hospitalization |
| Fatigue | No interference with activity | Some interference with activity | Significant; prevents daily activity | Requires emergency department visit or hospitalization |
| Myalgia (muscle aches all over body) | No interference with activity | Some interference with activity | Significant; prevents daily activity | Requires emergency department  visit or hospitalization |
| Arthralgia (joint aches in several joints) | No interference with activity | Some interference with activity | Significant; prevents daily activity | Requires emergency department visit or hospitalization |
| Nausea/vomiting | No interference with activity or 1 or 2 episodes/ 24 hours | Some interference with activity or  >2 episodes/ 24 hours | Prevents daily activity, requires outpatient intravenous hydration | Requires emergency department visit or hospitalization for hypotensive shock |
| Chills | No interference with activity | Some interference with activity not requiring medical intervention | Prevents daily activity and requires medical intervention | Requires emergency department  visit or hospitalization |
| Fever (oral) | 38.0°C to 38.4°C  100.4°F to 101.1°F | 38.5°C to 38.9°C  101.2°F to 102.0°F | 39.0°C to 40.0°C  102.1°F to 104.0°F | >40.0°C  >104.0°F |

AE, adverse event; eCRF, electronic case report form

Note: Events listed above but starting >7 days post-study injection will be recorded on the AE page of the eCRF. Causality for each event will be determined per assessment by the investigator.

**Analysis populations**

The safety set consisted of all randomly assigned participants who received a study vaccine. Participants were analyzed according to the study vaccine received. The solicited safety set consisted of all participants in the safety set who contributed any solicited adverse reaction data. The full analysis set consisted of all randomly assigned participants who received a study vaccine. Participants were analyzed according to which group they were randomized to. The per-protocol set consisted of all participants in the full analysis set who received a randomized study vaccine and had no major protocol deviations that impacted the immune response (eg, major deviations from the study injection schedule or timings of immunogenicity blood sampling).

**Supplementary Results**

**Immunogenicity**

*mRNA-1273.222*

Peak nAb GMCs (95% CIs) at Day 15 for the younger and older groups, respectively, were 10814.43 (8367.54, 13976.85) and 11370.47 (9480.98, 13636.51) for Ancetral; 3174.73 (2236.87, 4505.83) and 3137.28 (2231.64, 4410.44) for BA.1 (B.1.1.529); and 1687.82 (1197.06, 2379.77) and 1791.34 (1273.48, 2519.78) for the BA.4/5 variant. For all SARS-CoV-2 variants tested, boosting, as measured by GMFR at Day 29 in the total and seropositive populations, was similar between the 2 age groups, with the largest increases from baseline observed in SARS-CoV-2–seronegative versus seropositive participants (**Figure S3**). SRRs were higher for all 3 SARS-CoV-2 variants for participants who were SARS-CoV-2–seronegative versus participants who were SARS-CoV-2–seropositive at baseline Ancestral, 63.6% vs 42.6%; BA.1, 90.9% vs 57.4%; BA.4/5, 72.7% vs 55.3%; **Table S6**).

*mRNA-1345*

Peak GMTs (95% CIs) at Day 15 in the younger and older groups, respectively, were 19291.02 (13323.36, 27931.63) and 16454.63 (11261.48, 24042.56) against RSV-A, and 9269.48 (6764.10, 12702.84) and 7542.18 (5304.91, 10723.00) against RSV-B. GMTs (95% CIs) against RSV-A were higher than those against RSV-B. For RSV-A and RSV-B, boosting, as measured by GMFR, was generally higher in participants aged 18 to 49 years (RSV-A Day 8, Day 15, and Day 29: 6.5, 12.7, and 10.1, respectively; RSV-B Day 8, Day 15, and Day 29: 3.7, 7.3, and 5.0, respectively) compared with those aged 50 to 75 years (RSV-A Day 8, Day 15, and Day 29: 4.0, 7.0, and 6.3, respectively; RSV-B Day 8, Day 15, and Day 29: 3.4, 4.9, and 4.1, respectively) (**Figure 3**). Overall, the SRR (based on a 4-fold rise from baseline) was higher for RSV-A (71.4%) than for RSV-B (53.6%). SRRs were higher in adults aged 18 to 49 years than those aged 50 to 75 years for RSV-A (85.2% vs 58.6%) but were similar for RSV-B (51.9% vs 55.2%) at Day 29 (**Table S7**).

**Supplementary Figures**

**Figure S1. Participant disposition.**

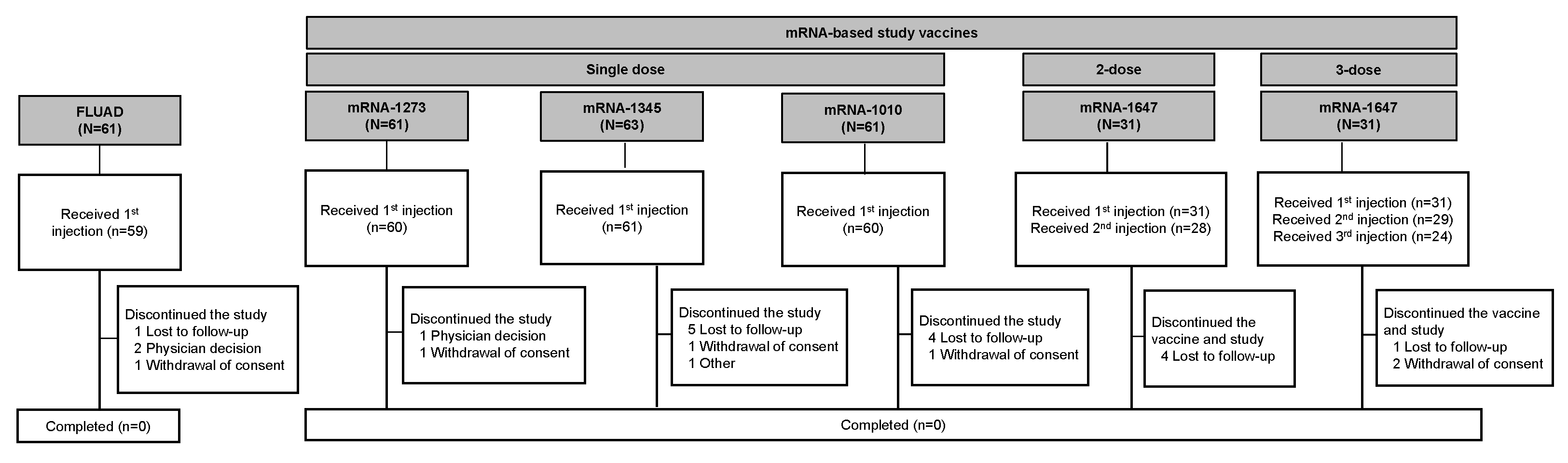

**Figure S2. Solicited local and systemic ARs through 7 days after mRNA-1647 by dose, grade, and baseline CMV serostatus for the 2- and 3-dose groups (first and second injection solicited safety set).**

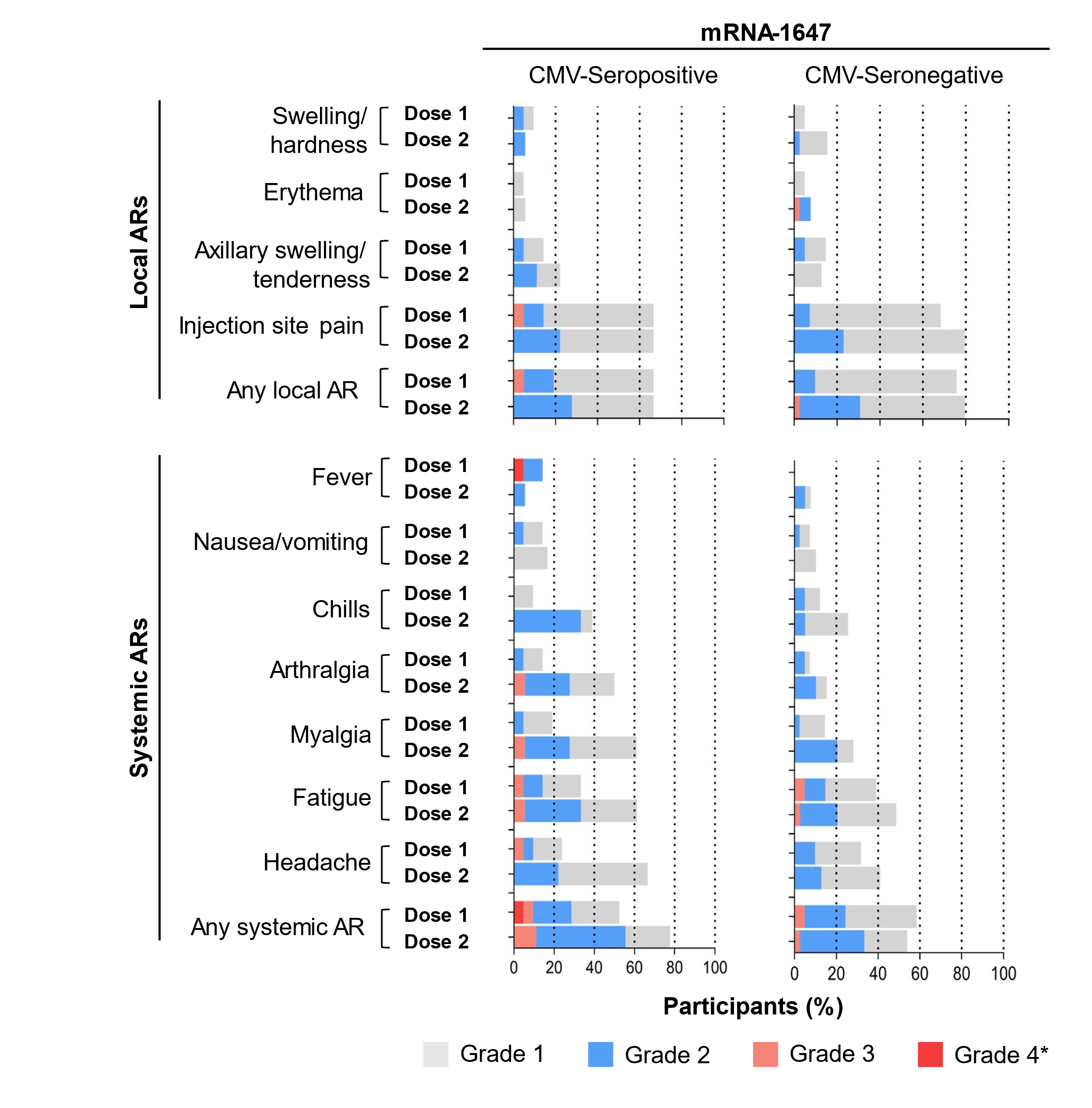

AR, adverse reaction; CMV, cytomegalovirus.

Data for 2- and 3-dose mRNA-1647 groups were combined and are shown by dose in the first injection solicited safety set (dose 1) and second injection solicited safety set (dose 2) for mRNA-1647. *All grade 4 events were fevers and were the result of self-reported data entry errors and are reported verbatim as recorded in the eDiary.

**Figure S3. Antibody levels for participants vaccinated with mRNA-1273.222 by SARS-CoV-2 serostatus (anti-nucleoprotein positive or negative) and age group (per-protocol set).**

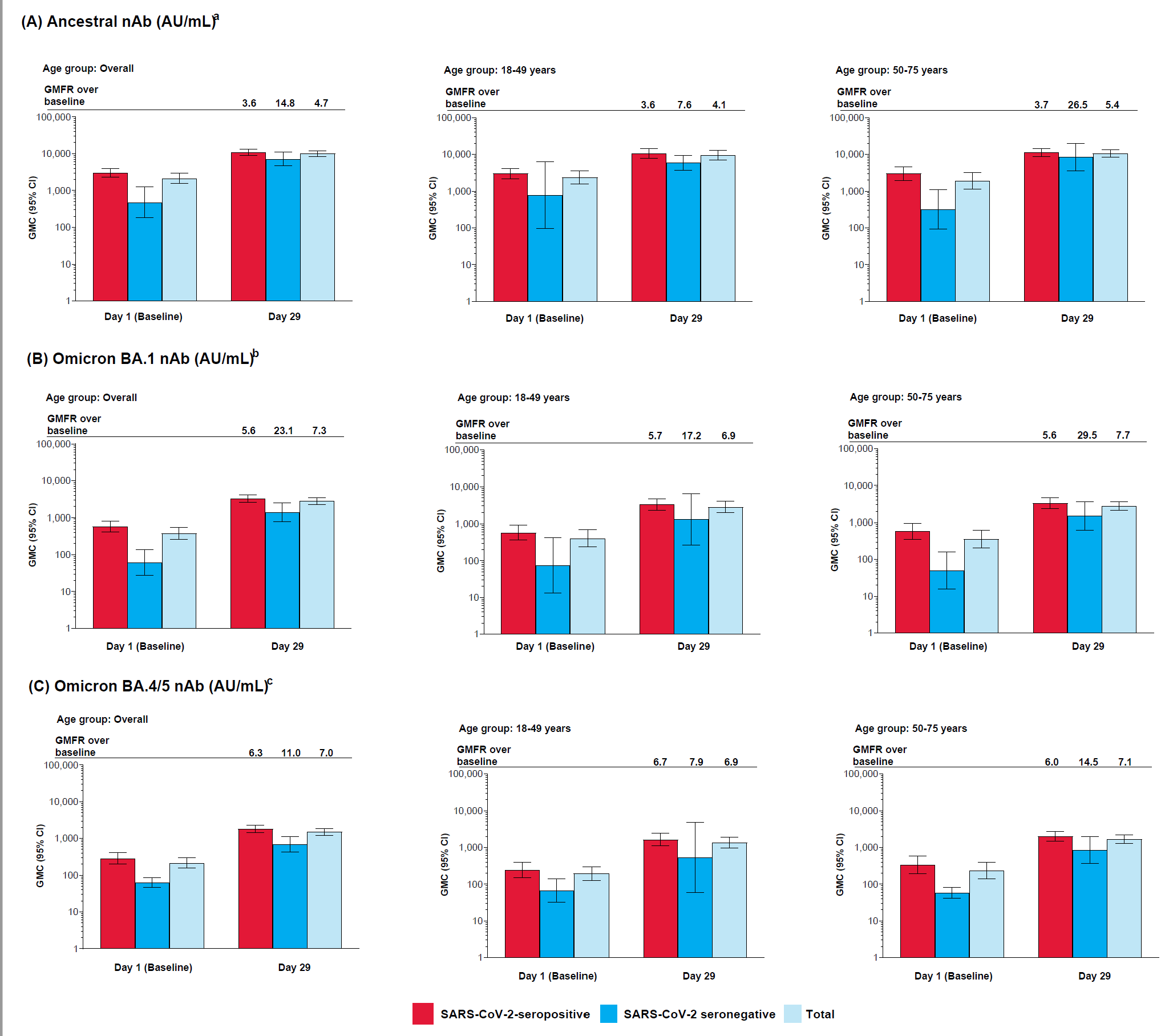

CI, confidence interval; GMFR, geometric mean fold rise, GMT, geometric mean titer; LLOQ, lower limit of quantification; nAb, neutralizing antibody; ULOQ, upper limit of quantification.

^a^LLOQ: 10; ULOQ: 111,433.

^b^LLOQ: 8; ULOQ: 41,984.

^c^LLOQ: 103; ULOQ: 28,571.

Antibody values reported as below the LLOQ are replaced by 0.5 × LLOQ. Values greater than the ULOQ are converted to the ULOQ.

COVID = SARS-CoV-2 serology sample test at baseline. Participants missing SARS-CoV-2 serology sample test results at baseline were excluded from the figure.

**Figure S4. Cellular immunogenicity against ancestral SARS-CoV-2 Ancestral and omicron-BA.4/5 following vaccination with mRNA-1273.222. (A) Overall T cell responses (age groups combined); (B) Th-1–specific T cell responses by age group.**

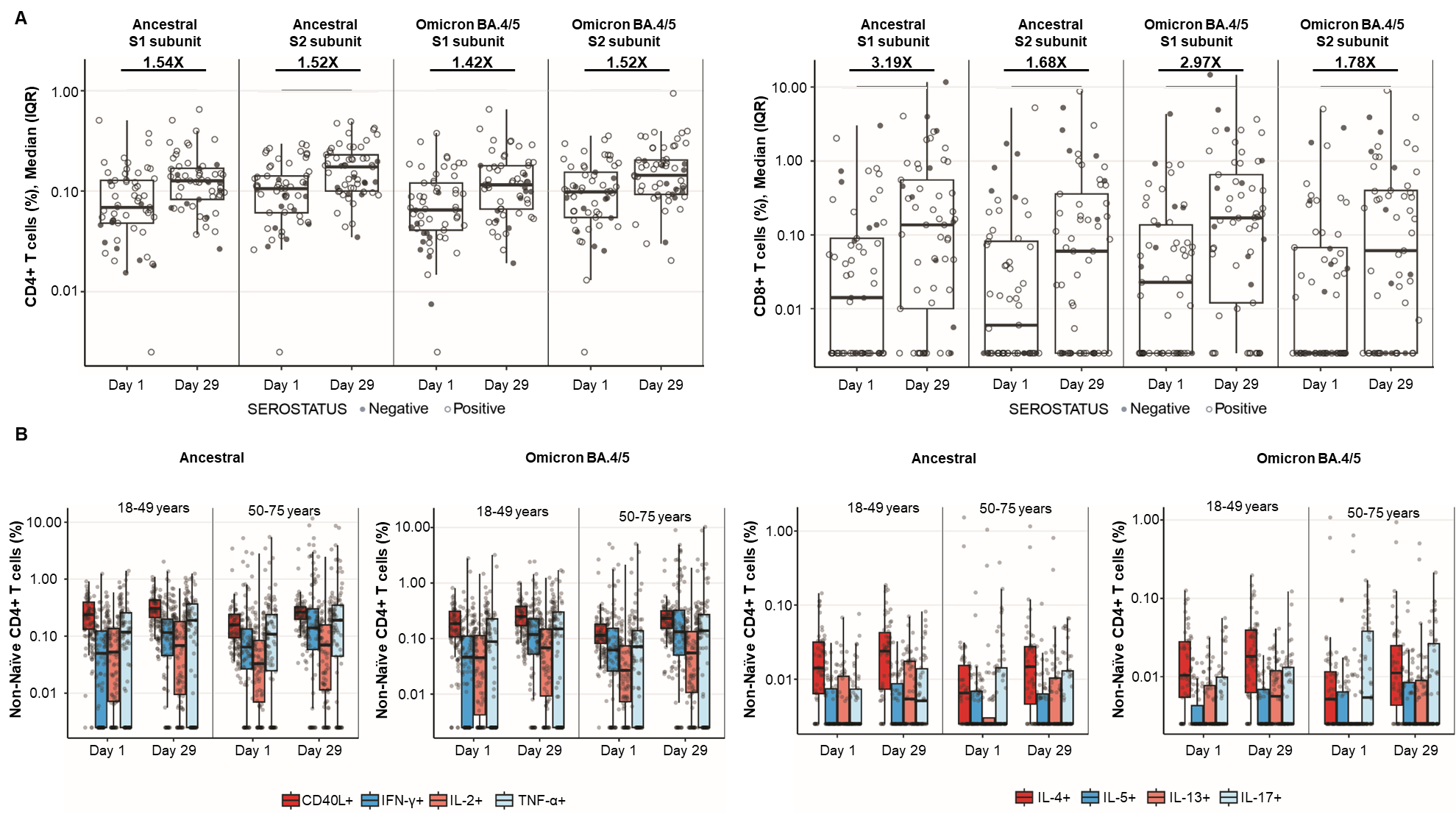

The solid black data points represent boxplot outliers.

**Figure S5. Percentage of IFN-γ plus RSV preF-specific CD4^+^ and CD8^+^ T cells following vaccination with mRNA-1345. (A) CD4^+^ or CD8^+^ T cells for the combined age cohorts; (B) CD4+ T cell GMFRs (95% CIs) by age cohorts^a^**

**A**

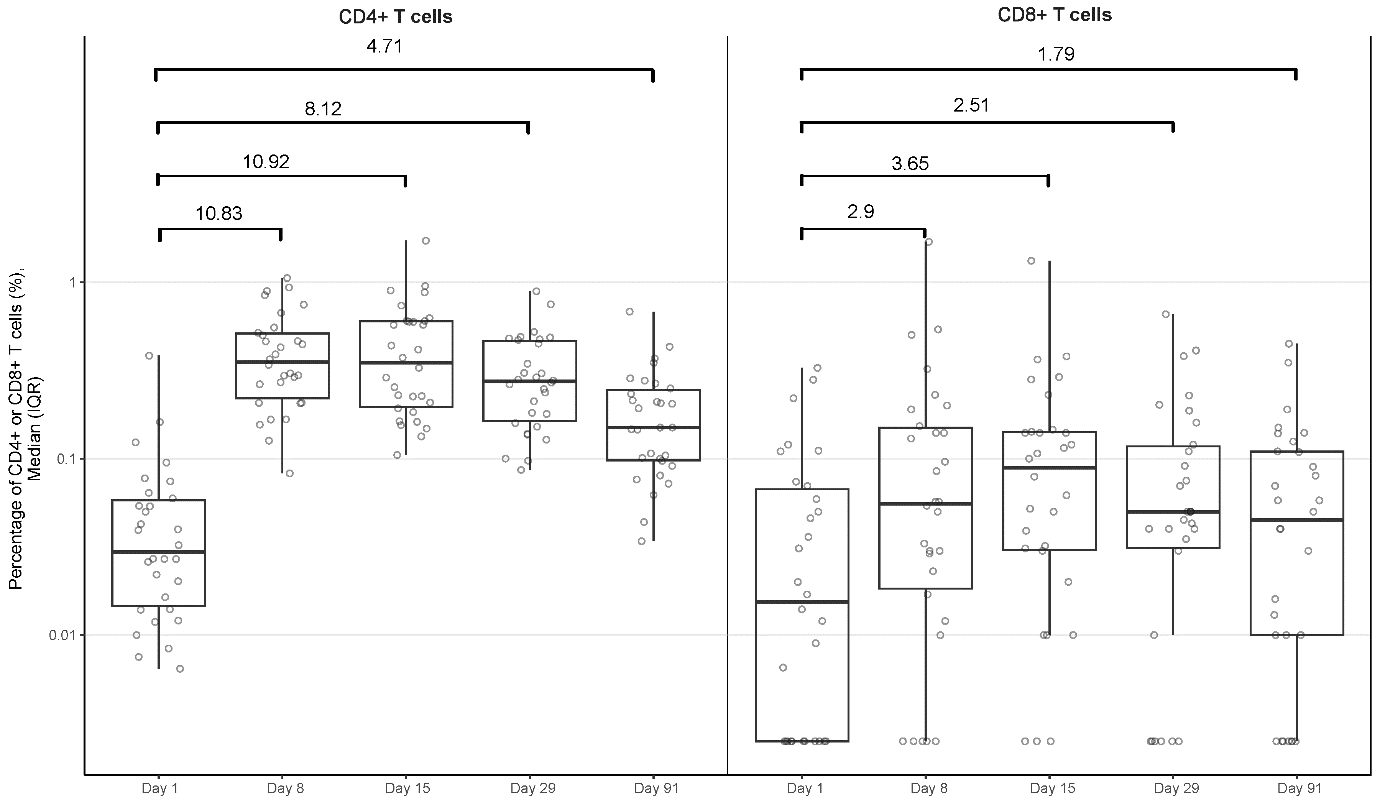

**B**

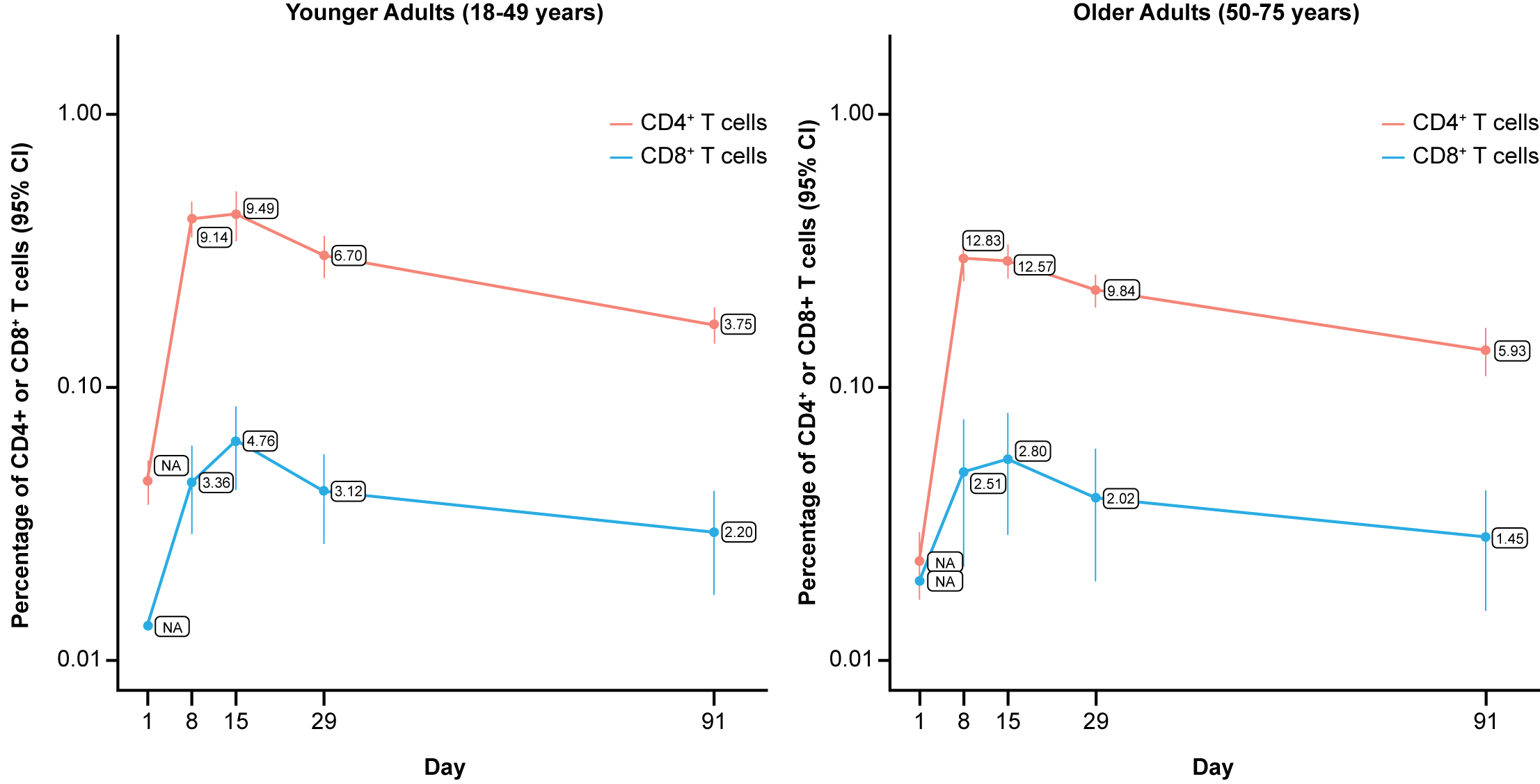

CI, confidence interval; GMFR, geometric mean fold rise; IFN-γ, interferon gamma; RSV, respiratory syncytial virus.

CD4^+^ and CD8^+^ T-cell subsets were gated on CD69^+^ and IFN-γ+.

**Figure S6. Geometric mean (95% CI) by vaccination group, study visit, and CMV serostatus (mRNA-1647 group) for (A) additional cytokine biomarkers and (B) selected cytokines/chemokines, which were not detected after vaccination.**

**A**

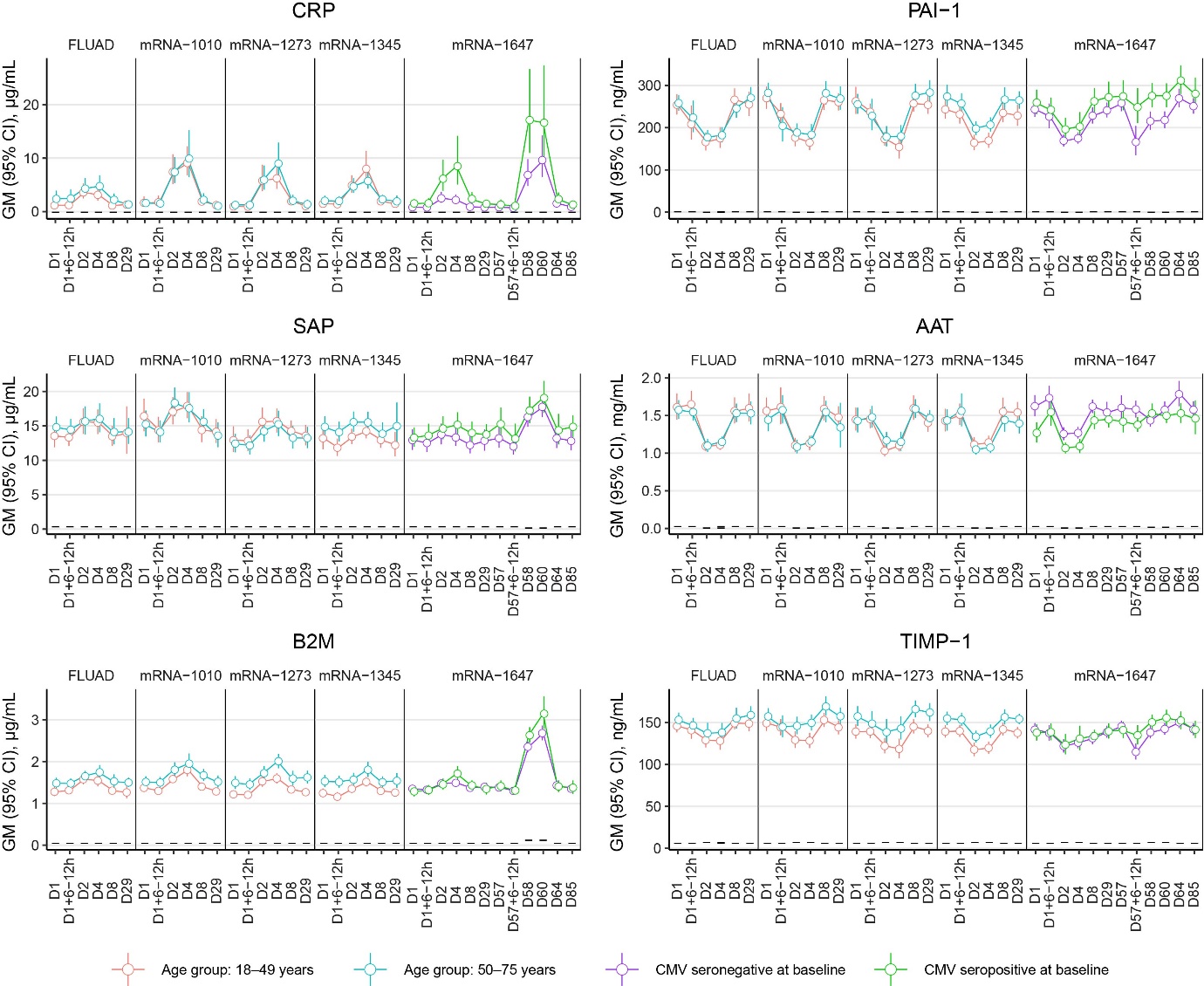

**B**

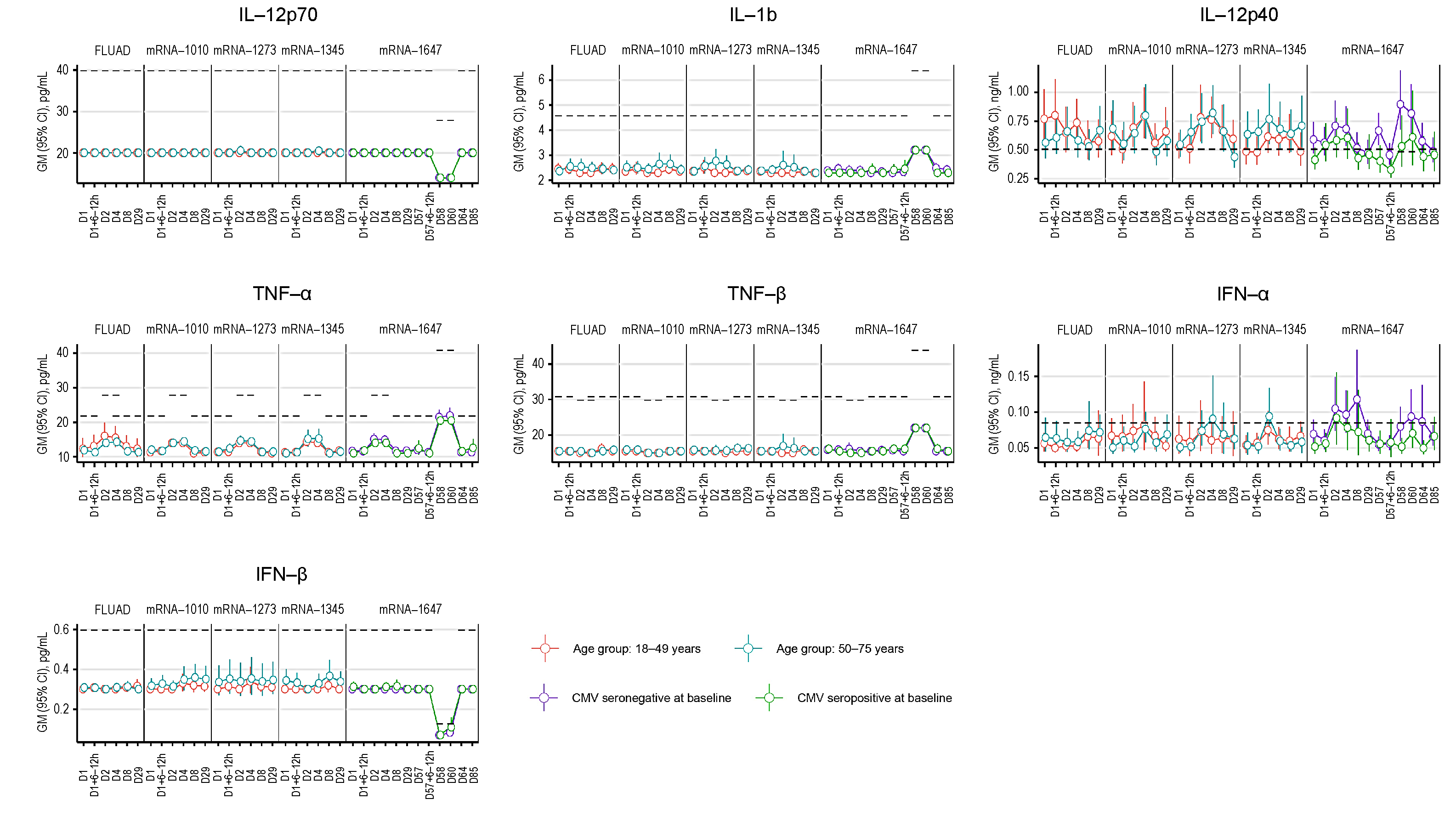

Dotted lines represent the LLOQ for each analyte. For some time points, the LLOQ is either decreased or increased, since not all samples could be assessed prior to the establishment of a revised LLOQ for samples run at a later time point.

AAT, alpha-1 antitrypsin, B2M, beta-2 macroglobulin; CRP, C-reactive protein;
IFN-α, interferon-α; IFN-β, interferon-β; IL-12p40, interleukin-12 subunit p40; IL-12p70, interleukin-12 subunit p70; IL-1β, interleukin-β; LLOQ, lower limit of quantification; PAI-1, plasminogen

activator inhibitor-1; SAP, serum amyloid P; TIMP-1, tissue inhibitor of metalloproteinase 1; TNF-α, tumor necrosis factor-α; TNF-β, tumor necrosis factor-β.

**Figure S7. Correlation between serum cytokine biomarker responses (normalized area under the curve) and reactogenicity sum (sum of all grades across symptoms) for the (A) mRNA groups and the (B) FLUAD group.**

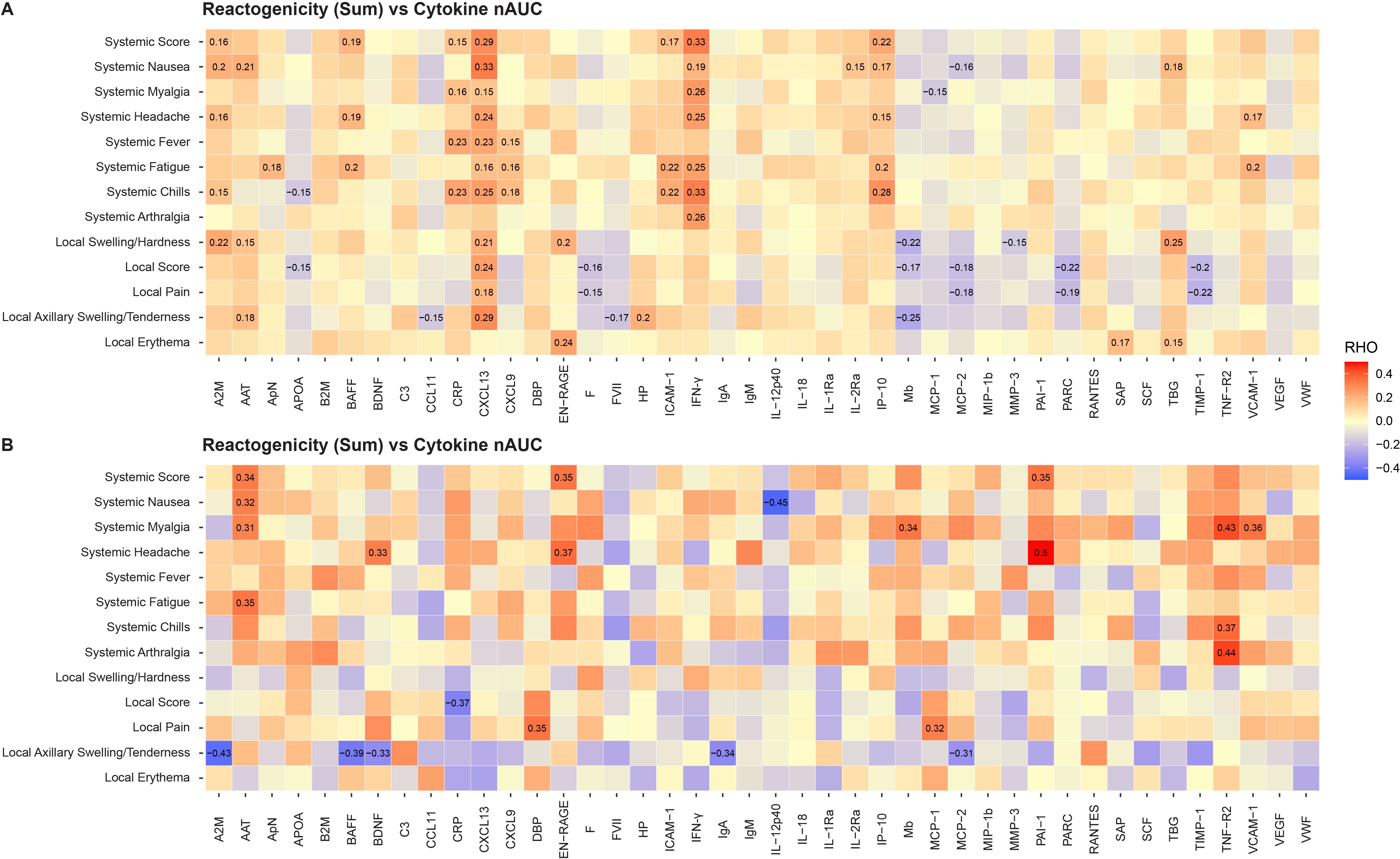

For all mRNA vaccine groups combined and the adjuvanted quadrivalent influenza vaccine (FLUAD). Statistically significant values are labeled with the correlation coefficient.

A2M, alpha-2-macroglobulin; AAT, alpha-1-antitrypsin; ApN, adiponectin; APOA, apolipoprotein(a); B2M, beta-2-microglobulin; BAFF, B-cell activating factor; BDNF, brain-derived neurotrophic factor; C3, complement C3; CCL11, C-C motif chemokine 11; CRP, C-reactive protein; CXCL9, CXC chemokine ligand 9; CXCL13, CXC chemokine ligand 13; DBP, vitamin D-binding protein; EN-RAGE, extracellular newly identified receptor for advanced glycation end products binding protein; F, ferritin; FVII, factor VII; HP, haptoglobin; ICAM-1, intercellular adhesion molecule 1; IFN-γ, interferon gamma; IgA, immunoglobulin A; IgM, immunoglobulin M; IL-12p40, interleukin-12 subunit p40; IL-18, interleukin-18; IL-1Ra, interleukin-1 receptor antagonist; IL-2Rα, interleukin-2 receptor α; IP-10, interferon gamma induced protein 10; Mb, myoglobin; MCP-1, monocyte chemotactic protein 1; MCP-2, monocyte chemotactic protein 2; MIP-1β, macrophage inflammatory protein-1 β; MMP-3, matrix metalloproteinase-3; PAI-1, plasminogen activator inhibitor 1; PARC, pulmonary and activation-regulated chemokine; RANTES, T-cell−specific protein RANTES; SAP, serum amyloid P-component; SCF, stem cell factor; TBG, thyroxine-binding globulin; TIMP-1, tissue inhibitor of metalloproteinases 1; TNF-R2, tumor necrosis factor receptor 2; VCAM-1, vascular cell adhesion molecule-1; VEGF, vascular endothelial growth factor; VWF, von Willebrand factor.

**Supplementary Tables**

**Supplementary Table 1. Solicited local and systemic ARs within 7 days after injection by grade and age group - first injection (solicited safety set)**

|  | **mRNA-1345**  **n=61** | | **mRNA-1273.222**  **n=60** | | **mRNA-1010**  **n=60** | | **FLUAD**  **n=59** | |
| --- | --- | --- | --- | --- | --- | --- | --- | --- |
|  | **18-49 years** | **50-75 years** | **18-49 years** | **50-75 years** | **18-49 years** | **50-75 years** | **18-49 years** | **50-75 years** |
| Solicited local ARs, n (%)^a^ | 32 | 29 | 29 | 31 | 30 | 30 | 29 | 30 |
| Any solicited local ARs, n (%)^b^ | 17 (53.1) | 17 (58.6) | 23 (79.3) | 20 (64.5) | 20 (66.7) | 17 (56.7) | 15 (51.7) | 18 (60.0) |
| 95% CI | 36.0, 72.7 | 38.9, 76.5 | 60.3, 92.0 | 47.2, 82.7 | 47.2, 82.7 | 37.4, 74.5 | 32.5, 70.6 | 40.6, 77.3 |
| Grade 1 | 13 (40.6) | 13 (44.8) | 20 (69.0) | 17 (54.8) | 11 (36.7) | 13 (43.3) | 14 (48.3) | 15 (50.0) |
| Grade 2 | 4 (12.5) | 4 (13.8) | 3 (10.3) | 3 (9.7) | 9 (30.0) | 4 (13.3) | 1 (3.4) | 2 (6.7) |
| Grade 3 | 0 | 0 | 0 | 0 | 0 | 0 | 0 | 1 (3.3) |
| Grade 4 | 0 | 0 | 0 | 0 | 0 | 0 | 0 | 0 |
| Grade 3 or grade 4 | 0 | 0 | 0 | 0 | 0 | 0 | 0 | 1 (3.3) |
| Injection site pain | 32 | 29 | 29 | 31 | 30 | 30 | 29 | 30 |
| Any | 17 (53.1) | 17 (58.6) | 23 (79.3) | 20 (64.5) | 19 (63.3) | 17 (56.7) | 13 (44.8) | 17 (56.7) |
| Grade 1 | 13 (40.6) | 13 (44.8) | 20 (69.0) | 17 (54.8) | 13 (43.3) | 13 (43.3) | 13 (44.8) | 14 (46.7) |
| Grade 2 | 4 (12.5) | 4 (13.8) | 3 (10.3) | 3 (9.7) | 6 (20.0) | 4 (13.3) | 0 | 2 (6.7) |
| Grade 3 | 0 | 0 | 0 | 0 | 0 | 0 | 0 | 1 (3.3) |
| Grade 4 | 0 | 0 | 0 | 0 | 0 | 0 | 0 | 0 |
| Grade 3 or grade 4 | 0 | 0 | 0 | 0 | 0 | 0 | 0 | 1 (3.3) |
| Erythema (redness) | 32 | 29 | 29 | 31 | 30 | 30 | 29 | 30 |
| Any | 0 | 0 | 1 (3.4) | 1 (3.2) | 0 | 0 | 1 (3.4) | 0 |
| Grade 1 | 0 | 0 | 1 (3.4) | 1 (3.2) | 0 | 0 | 0 | 0 |
| Grade 2 | 0 | 0 | 0 | 0 | 0 | 0 | 1 (3.4) | 0 |
| Grade 3 | 0 | 0 | 0 | 0 | 0 | 0 | 0 | 0 |
| Grade 4 | 0 | 0 | 0 | 0 | 0 | 0 | 0 | 0 |
| Grade 3 or grade 4 | 0 | 0 | 0 | 0 | 0 | 0 | 0 | 0 |
| Swelling/induration (hardness) | 32 | 29 | 29 | 31 | 30 | 30 | 29 | 30 |
| Any | 0 | 0 | 1 (3.4) | 0 | 1 (3.3) | 0 | 0 | 1 (3.3) |
| Grade 1 | 0 | 0 | 1 (3.4) | 0 | 0 | 0 | 0 | 1 (3.3) |
| Grade 2 | 0 | 0 | 0 | 0 | 1 (3.3) | 0 | 0 | 0 |
| Grade 3 | 0 | 0 | 0 | 0 | 0 | 0 | 0 | 0 |
| Grade 4 | 0 | 0 | 0 | 0 | 0 | 0 | 0 | 0 |
| Grade 3 or grade 4 | 0 | 0 | 0 | 0 | 0 | 0 | 0 | 0 |
| Axillary (underarm) swelling or tenderness | 32 | 29 | 29 | 31 | 30 | 30 | 29 | 30 |
| Any | 7 (21.9) | 5 (17.2) | 5 (17.2) | 3 (9.7) | 9 (30.0) | 1 (3.3) | 2 (6.9) | 3 (10.0) |
| Grade 1 | 6 (18.8) | 5 (17.2) | 5 (17.2) | 3 (9.7) | 6 (20.0) | 0 | 2 (6.9) | 3 (10.0) |
| Grade 2 | 1 (3.1) | 0 | 0 | 0 | 3 (10.0) | 1 (3.3) | 0 | 0 |
| Grade 3 | 0 | 0 | 0 | 0 | 0 | 0 | 0 | 0 |
| Grade 4 | 0 | 0 | 0 | 0 | 0 | 0 | 0 | 0 |
| Grade 3 or grade 4 | 0 | 0 | 0 | 0 | 0 | 0 | 0 | 0 |
| Solicited systemic ARs, n (%)^a^ | 32 | 29 | 29 | 31 | 30 | 30 | 29 | 30 |
| Any solicited systemic ARs, n (%)^b^ | 12 (37.5) | 14 (48.3) | 18 (62.1) | 19 (61.3) | 18 (60.0) | 16 (53.3) | 13 (44.8) | 13 (43.3) |
| 95% CI | 21.8, 57.8 | 29.4, 67.5 | 42.3, 79.3 | 43.9, 80.1 | 40.6, 77.3 | 34.3, 71.7 | 26.4, 64.3 | 25.5, 62.6 |
| Grade 1 | 5 (15.6) | 8 (27.6) | 10 (34.5) | 11 (35.5) | 5 (16.7) | 8 (26.7) | 9 (31.0) | 7 (23.3) |
| Grade 2 | 6 (18.8) | 5 (17.2) | 6 (20.7) | 8 (25.8) | 12 (40.0) | 7 (23.3) | 3 (10.3) | 3 (10.0) |
| Grade 3 | 0 | 0 | 2 (6.9) | 0 | 1 (3.3) | 1 (3.3) | 1 (3.4) | 3 (10.0) |
| Grade 4 | 1 (3.1) | 1 (3.4) | 0 | 0 | 0 | 0 | 0 | 0 |
| Grade 3 or grade 4 | 1 (3.1) | 1 (3.4) | 2 (6.9) | 0 | 1 (3.3) | 1 (3.3) | 1 (3.4) | 3 (10.0) |
| Fever | 32 | 29 | 29 | 31 | 30 | 30 | 29 | 30 |
| Any^c^ | 3 (9.4) | 1 (3.4) | 0 | 1 (3.2) | 2 (6.7) | 2 (6.7) | 0 | 2 (6.7) |
| Grade 1 | 1 (3.1) | 0 | 0 | 1 (3.2) | 2 (6.7) | 2 (6.7) | 0 | 2 (6.7) |
| Grade 2 | 1 (3.1) | 0 | 0 | 0 | 0 | 0 | 0 | 0 |
| Grade 3 | 0 | 0 | 0 | 0 | 0 | 0 | 0 | 0 |
| Grade 4^c^ | 1 (3.1) | 1 (3.4) | 0 | 0 | 0 | 0 | 0 | 0 |
| Grade 3 or grade 4 | 1 (3.1) | 1 (3.4) | 0 | 0 | 0 | 0 | 0 | 0 |
| Headache | 32 | 29 | 29 | 31 | 30 | 30 | 29 | 30 |
| Any | 5 (15.6) | 6 (20.7) | 15 (51.7) | 12 (38.7) | 9 (30.0) | 9 (30.0) | 9 (31.0) | 11 (36.7) |
| Grade 1 | 3 (9.4) | 5 (17.2) | 10 (34.5) | 10 (32.3) | 5 (16.7) | 7 (23.3) | 8 (27.6) | 7 (23.3) |
| Grade 2 | 2 (6.3) | 1 (3.4) | 3 (10.3) | 2 (6.5) | 4 (13.3) | 2 (6.7) | 0 | 3 (10.0) |
| Grade 3 | 0 | 0 | 2 (6.9) | 0 | 0 | 0 | 1 (3.4) | 1 (3.3) |
| Grade 4 | 0 | 0 | 0 | 0 | 0 | 0 | 0 | 0 |
| Grade 3 or grade 4 | 0 | 0 | 2 (6.9) | 0 | 0 | 0 | 1 (3.4) | 1 (3.3) |
| Fatigue | 32 | 29 | 29 | 31 | 30 | 30 | 29 | 30 |
| Any | 8 (25.0) | 10 (34.5) | 12 (41.4) | 13 (41.9) | 11 (36.7) | 14 (46.7) | 9 (31.0) | 10 (33.3) |
| Grade 1 | 5 (15.6) | 6 (20.7) | 6 (20.7) | 10 (32.3) | 4 (13.3) | 8 (26.7) | 5 (17.2) | 4 (13.3) |
| Grade 2 | 3 (9.4) | 4 (13.8) | 5 (17.2) | 3 (9.7) | 7 (23.3) | 5 (16.7) | 3 (10.3) | 5 (16.7) |
| Grade 3 | 0 | 0 | 1 (3.4) | 0 | 0 | 1 (3.3) | 1 (3.4) | 1 (3.3) |
| Grade 4 | 0 | 0 | 0 | 0 | 0 | 0 | 0 | 0 |
| Grade 3 or grade 4 | 0 | 0 | 1 (3.4) | 0 | 0 | 1 (3.3) | 1 (3.4) | 1 (3.3) |
| Myalgia | 32 | 29 | 29 | 31 | 30 | 30 | 29 | 30 |
| Any | 7 (21.9) | 6 (20.7) | 11 (37.9) | 11 (35.5) | 13 (43.3) | 7 (23.3) | 5 (17.2) | 9 (30.0) |
| Grade 1 | 3 (9.4) | 3 (10.3) | 8 (27.6) | 6 (19.4) | 5 (16.7) | 3 (10.0) | 3 (10.3) | 5 (16.7) |
| Grade 2 | 4 (12.5) | 3 (10.3) | 2 (6.9) | 5 (16.1) | 7 (23.3) | 4 (13.3) | 2 (6.9) | 3 (10.0) |
| Grade 3 | 0 | 0 | 1 (3.4) | 0 | 1 (3.3) | 0 | 0 | 1 (3.3) |
| Grade 4 | 0 | 0 | 0 | 0 | 0 | 0 | 0 | 0 |
| Grade 3 or grade 4 | 0 | 0 | 1 (3.4) | 0 | 1 (3.3) | 0 | 0 | 1 (3.3) |
| Arthralgia | 32 | 29 | 29 | 31 | 30 | 30 | 29 | 30 |
| Any | 5 (15.6) | 4 (13.8) | 10 (34.5) | 5 (16.1) | 8 (26.7) | 6 (20.0) | 1 (3.4) | 6 (20.0) |
| Grade 1 | 2 (6.3) | 3 (10.3) | 9 (31.0) | 3 (9.7) | 4 (13.3) | 3 (10.0) | 1 (3.4) | 1 (3.3) |
| Grade 2 | 3 (9.4) | 1 (3.4) | 1 (3.4) | 2 (6.5) | 3 (10.0) | 3 (10.0) | 0 | 4 (13.3) |
| Grade 3 | 0 | 0 | 0 | 0 | 1 (3.3) | 0 | 0 | 1 (3.3) |
| Grade 4 | 0 | 0 | 0 | 0 | 0 | 0 | 0 | 0 |
| Grade 3 or grade 4 | 0 | 0 | 0 | 0 | 1 (3.3) | 0 | 0 | 1 (3.3) |
| Nausea/vomiting | 32 | 29 | 29 | 31 | 30 | 30 | 29 | 30 |
| Any | 2 (6.3) | 1 (3.4) | 5 (17.2) | 3 (9.7) | 2 (6.7) | 0 | 1 (3.4) | 3 (10.0) |
| Grade 1 | 2 (6.3) | 1 (3.4) | 4 (13.8) | 2 (6.5) | 2 (6.7) | 0 | 0 | 2 (6.7) |
| Grade 2 | 0 | 0 | 1 (3.4) | 1 (3.2) | 0 | 0 | 1 (3.4) | 1 (3.3) |
| Grade 3 | 0 | 0 | 0 | 0 | 0 | 0 | 0 | 0 |
| Grade 4 | 0 | 0 | 0 | 0 | 0 | 0 | 0 | 0 |
| Grade 3 or grade 4 | 0 | 0 | 0 | 0 | 0 | 0 | 0 | 0 |
| Chills | 32 | 29 | 29 | 31 | 30 | 30 | 29 | 30 |
| Any | 4 (12.5) | 1 (3.4) | 4 (13.8) | 8 (25.8) | 9 (30.0) | 5 (16.7) | 3 (10.3) | 6 (20.0) |
| Grade 1 | 2 (6.3) | 0 | 2 (6.9) | 5 (16.1) | 2 (6.7) | 5 (16.7) | 1 (3.4) | 3 (10.0) |
| Grade 2 | 2 (6.3) | 1 (3.4) | 2 (6.9) | 3 (9.7) | 7 (23.3) | 0 | 2 (6.9) | 2 (6.7) |
| Grade 3 | 0 | 0 | 0 | 0 | 0 | 0 | 0 | 1 (3.3) |
| Grade 4 | 0 | 0 | 0 | 0 | 0 | 0 | 0 | 0 |
| Grade 3 or grade 4 | 0 | 0 | 0 | 0 | 0 | 0 | 0 | 1 (3.3) |

AR, adverse reaction; CI, confidence interval.

^a^Number of exposed subjects who submitted any data for the event.

^b^Percentages are based on the number of exposed subjects who submitted any data for the event.

^c^All Grade 4 fevers were the result of self-reported data entry errors.

Any = grade 1 or higher.

**Supplementary Table 2. Solicited local and systemic ARs within 7 days after injection by grade and baseline CMV serostatus for 2- and 3-dose groups (mRNA-1647) (first and second injection solicited safety set)**

|  | **mRNA-1647 Total^a^** | | | | | |
| --- | --- | --- | --- | --- | --- | --- |
|  | **First Injection** | | | **Second Injection** | | |
|  | **CMV Positive**  **n=21** | **CMV Negative**  **n=41** | **Total**  **n=62** | **CMV Positive**  **n=18** | **CMV Negative**  **n=39** | **Total**  **n=57** |
| Solicited local ARs, n (%)^b^ | 21 | 41 | 62 | 18 | 39 | 57 |
| Any solicited local ARs, n (%)^c^ | 14 (66.7) | 31 (75.6) | 45 (72.6) | 12 (66.7) | 31 (79.5) | 43 (75.4) |
| 95% CI | 43.0, 85.4 | 59.7, 87.6 | 59.8, 83.1 | 41.0, 86.7 | 63.5, 90.7 | 62.2, 85.9 |
| Grade 1 | 10 (47.6) | 27 (65.9) | 37 (59.7) | 7 (38.9) | 19 (48.7) | 26 (45.6) |
| Grade 2 | 3 (14.3) | 4 (9.8) | 7 (11.3) | 5 (27.8) | 11 (28.2) | 16 (28.1) |
| Grade 3 | 1 (4.8) | 0 | 1 (1.6) | 0 | 1 (2.6) | 1 (1.8) |
| Grade 4 | 0 | 0 | 0 | 0 | 0 | 0 |
| Grade 3 or 4 | 1 (4.8) | 0 | 1 (1.6) | 0 | 1 (2.6) | 1 (1.8) |
| Injection site ain | 21 | 41 | 62 | 18 | 39 | 57 |
| Any | 14 (66.7) | 28 (68.3) | 42 (67.7) | 12 (66.7) | 31 (79.5) | 43 (75.4) |
| Grade 1 | 11 (52.4) | 25 (61.0) | 36 (58.1) | 8 (44.4) | 22 (56.4) | 30 (52.6) |
| Grade 2 | 2 (9.5) | 3 (7.3) | 5 (8.1) | 4 (22.2) | 9 (23.1) | 13 (22.8) |
| Grade 3 | 1 (4.8) | 0 | 1 (1.6) | 0 | 0 | 0 |
| Grade 4 | 0 | 0 | 0 | 0 | 0 | 0 |
| Grade 3 or 4 | 1 (4.8) | 0 | 1 (1.6) | 0 | 0 | 0 |
| Erythema (redness) | 21 | 41 | 62 | 18 | 39 | 57 |
| Any | 1 (4.8) | 2 (4.9) | 3 (4.8) | 1 (5.6) | 3 (7.7) | 4 (7.0) |
| Grade 1 | 1 (4.8) | 2 (4.9) | 3 (4.8) | 1 (5.6) | 0 | 1 (1.8) |
| Grade 2 | 0 | 0 | 0 | 0 | 2 (5.1) | 2 (3.5) |
| Grade 3 | 0 | 0 | 0 | 0 | 1 (2.6) | 1 (1.8) |
| Grade 4 | 0 | 0 | 0 | 0 | 0 | 0 |
| Grade 3 or 4 | 0 | 0 | 0 | 0 | 1 (2.6) | 1 (1.8) |
| Swelling/induration (hardness) | 21 | 41 | 62 | 18 | 39 | 57 |
| Any | 2 (9.5) | 2 (4.9) | 4 (6.5) | 1 (5.6) | 6 (15.4) | 7 (12.3) |
| Grade 1 | 1 (4.8) | 2 (4.9) | 3 (4.8) | 0 | 5 (12.8) | 5 (8.8) |
| Grade 2 | 1 (4.8) | 0 | 1 (1.6) | 1 (5.6) | 1 (2.6) | 2 (3.5) |
| Grade 3 | 0 | 0 | 0 | 0 | 0 | 0 |
| Grade 4 | 0 | 0 | 0 | 0 | 0 | 0 |
| Grade 3 or 4 | 0 | 0 | 0 | 0 | 0 | 0 |
| Axillary (underarm) swelling or tenderness | 21 | 41 | 62 | 18 | 39 | 57 |
| Any | 3 (14.3) | 6 (14.6) | 9 (14.5) | 4 (22.2) | 5 (12.8) | 9 (15.8) |
| Grade 1 | 2 (9.5) | 4 (9.8) | 6 (9.7) | 2 (11.1) | 5 (12.8) | 7 (12.3) |
| Grade 2 | 1 (4.8) | 2 (4.9) | 3 (4.8) | 2 (11.1) | 0 | 2 (3.5) |
| Grade 3 | 0 | 0 | 0 | 0 | 0 | 0 |
| Grade 4 | 0 | 0 | 0 | 0 | 0 | 0 |
| Grade 3 or 4 | 0 | 0 | 0 | 0 | 0 | 0 |
| Solicited systemic ARs | 21 | 41 | 62 | 18 | 39 | 57 |
| Any solicited systemic ARs | 11 (52.4) | 24 (58.5) | 35 (56.5) | 14 (77.8) | 21 (53.8) | 35 (61.4) |
| 95% CI | 29.8, 74.3 | 42.1, 73.7 | 43.3, 69.0 | 52.4, 93.6 | 37.2, 69.9 | 47.6, 74.0 |
| Grade 1 | 5 (23.8) | 14 (34.1) | 19 (30.6) | 4 (22.2) | 8 (20.5) | 12 (21.1) |
| Grade 2 | 4 (19.0) | 8 (19.5) | 12 (19.4) | 8 (44.4) | 12 (30.8) | 20 (35.1) |
| Grade 3 | 1 (4.8) | 2 (4.9) | 3 (4.8) | 2 (11.1) | 1 (2.6) | 3 (5.3) |
| Grade 4^d^ | 1 (4.8) | 0 | 1 (1.6) | 0 | 0 | 0 |
| Grade 3 or 4^d^ | 2 (9.5) | 2 (4.9) | 4 (6.5) | 2 (11.1) | 1 (2.6) | 3 (5.3) |
| Fever | 21 | 41 | 62 | 18 | 39 | 57 |
| Any | 3 (14.3) | 0 | 3 (4.8) | 1 (5.6) | 3 (7.7) | 4 (7.0) |
| Grade 1 | 0 | 0 | 0 | 0 | 1 (2.6) | 1 (1.8) |
| Grade 2 | 2 (9.5) | 0 | 2 (3.2) | 1 (5.6) | 2 (5.1) | 3 (5.3) |
| Grade 3 | 0 | 0 | 0 | 0 | 0 | 0 |
| Grade 4^d^ | 1 (4.8) | 0 | 1 (1.6) | 0 | 0 | 0 |
| Grade 3 or 4^d^ | 1 (4.8) | 0 | 1 (1.6) | 0 | 0 | 0 |
| Headache | 21 | 41 | 62 | 18 | 39 | 57 |
| Any | 5 (23.8) | 13 (31.7) | 18 (29.0) | 12 (66.7) | 16 (41.0) | 28 (49.1) |
| Grade 1 | 3 (14.3) | 9 (22.0) | 12 (19.4) | 8 (44.4) | 11 (28.2) | 19 (33.3) |
| Grade 2 | 1 (4.8) | 4 (9.8) | 5 (8.1) | 4 (22.2) | 5 (12.8) | 9 (15.8) |
| Grade 3 | 1 (4.8) | 0 | 1 (1.6) | 0 | 0 | 0 |
| Grade 4 | 0 | 0 | 0 | 0 | 0 | 0 |
| Grade 3 or 4 | 1 (4.8) | 0 | 1 (1.6) | 0 | 0 | 0 |
| Fatigue | 21 | 41 | 62 | 18 | 39 | 57 |
| Any | 7 (33.3) | 16 (39.0) | 23 (37.1) | 11 (61.1) | 19 (48.7) | 30 (52.6) |
| Grade 1 | 4 (19.0) | 10 (24.4) | 14 (22.6) | 5 (27.8) | 11 (28.2) | 16 (28.1) |
| Grade 2 | 2 (9.5) | 4 (9.8) | 6 (9.7) | 5 (27.8) | 7 (17.9) | 12 (21.1) |
| Grade 3 | 1 (4.8) | 2 (4.9) | 3 (4.8) | 1 (5.6) | 1 (2.6) | 2 (3.5) |
| Grade 4 | 0 | 0 | 0 | 0 | 0 | 0 |
| Grade 3 or 4 | 1 (4.8) | 2 (4.9) | 3 (4.8) | 1 (5.6) | 1 (2.6) | 2 (3.5) |
| Myalgia | 21 | 41 | 62 | 18 | 39 | 57 |
| Any | 4 (19.0) | 6 (14.6) | 10 (16.1) | 11 (61.1) | 11 (28.2) | 22 (38.6) |
| Grade 1 | 3 (14.3) | 5 (12.2) | 8 (12.9) | 6 (33.3) | 3 (7.7) | 9 (15.8) |
| Grade 2 | 1 (4.8) | 1 (2.4) | 2 (3.2) | 4 (22.2) | 8 (20.5) | 12 (21.1) |
| Grade 3 | 0 | 0 | 0 | 1 (5.6) | 0 | 1 (1.8) |
| Grade 4 | 0 | 0 | 0 | 0 | 0 | 0 |
| Grade 3 or 4 | 0 | 0 | 0 | 1 (5.6) | 0 | 1 (1.8) |
| Arthralgia | 21 | 41 | 62 | 18 | 39 | 57 |
| Any | 3 (14.3) | 3 (7.3) | 6 (9.7) | 9 (50.0) | 6 (15.4) | 15 (26.3) |
| Grade 1 | 2 (9.5) | 1 (2.4) | 3 (4.8) | 4 (22.2) | 2 (5.1) | 6 (10.5) |
| Grade 2 | 1 (4.8) | 2 (4.9) | 3 (4.8) | 4 (22.2) | 4 (10.3) | 8 (14.0) |
| Grade 3 | 0 | 0 | 0 | 1 (5.6) | 0 | 1 (1.8) |
| Grade 4 | 0 | 0 | 0 | 0 | 0 | 0 |
| Grade 3 or 4 | 0 | 0 | 0 | 1 (5.6) | 0 | 1 (1.8) |
| Nausea/vomiting | 21 | 41 | 62 | 18 | 39 | 57 |
| Any | 3 (14.3) | 3 (7.3) | 6 (9.7) | 3 (16.7) | 4 (10.3) | 7 (12.3) |
| Grade 1 | 2 (9.5) | 2 (4.9) | 4 (6.5) | 3 (16.7) | 4 (10.3) | 7 (12.3) |
| Grade 2 | 1 (4.8) | 1 (2.4) | 2 (3.2) | 0 | 0 | 0 |
| Grade 3 | 0 | 0 | 0 | 0 | 0 | 0 |
| Grade 4 | 0 | 0 | 0 | 0 | 0 | 0 |
| Grade 3 or 4 | 0 | 0 | 0 | 0 | 0 | 0 |
| Chills | 21 | 41 | 62 | 18 | 39 | 57 |
| Any | 2 (9.5) | 5 (12.2) | 7 (11.3) | 7 (38.9) | 10 (25.6) | 17 (29.8) |
| Grade 1 | 2 (9.5) | 3 (7.3) | 5 (8.1) | 1 (5.6) | 8 (20.5) | 9 (15.8) |
| Grade 2 | 0 | 2 (4.9) | 2 (3.2) | 6 (33.3) | 2 (5.1) | 8 (14.0) |
| Grade 3 | 0 | 0 | 0 | 0 | 0 | 0 |
| Grade 4 | 0 | 0 | 0 | 0 | 0 | 0 |
| Grade 3 or 4 | 0 | 0 | 0 | 0 | 0 | 0 |

AR, adverse reaction; CI, confidence interval; CMV, cytomegalovirus

^a^Data presented are those from the 2- and 3-dose groups combined (ie, total mRNA-1647 group).

^b^Number of exposed participants who submitted any data for the event.

^c^Percentages are based on the number of exposed participants who submitted any data for the event.

^d^All grade 4 fevers were the result of self-reported data entry errors.

Any = grade 1 or higher.

CMV serology results (positive or negative) are based on CMV prior to study treatment.

**Supplementary Table 3. Characteristics of solicited local and systemic ARs within 7 days after the first vaccine injection (first injection solicited safety set)**

|  | **mRNA-1273.222**  **n=60** | **mRNA-1345**  **n=61** | **Total mRNA-1647^a^**  **n=62** | **mRNA-1010**  **n=60** | **FLUAD**  **n=59** |
| --- | --- | --- | --- | --- | --- |
| Solicited local ARs^b^ | 59 | 60 | 62 | 60 | 59 |
| Any solicited local AR, n (%)^c^ | 43 (72.9) | 34 (56.7) | 45 (72.6) | 37 (61.7) | 33 (55.9) |
| Day of onset, median (min, max) | 1.0 (1, 2) | 1.0 (1, 2) | 1.0 (1, 3) | 1.0 (1, 3) | 1.0 (1, 7) |
| Duration^d^ (days), median (min, max) | 2.0 (1, 5) | 2.0 (1, 7) | 3.0 (1, 7) | 3.0 (1, 7) | 1.0 (1, 5) |
| Persisted beyond 7 days | 0 | 0 | 4 (6.5) | 0 | 0 |
| Solicited systemic ARs^b^ | 59 | 60 | 62 | 60 | 59 |
| Any solicited systemic AR, n (%)^c^ | 37 (62.7) | 26 (43.3) | 35 (56.5) | 34 (56.7) | 26 (44.1) |
| Day of onset, median (min, max) | 2.0 (1, 5) | 2.0 (1, 6) | 2.0 (1, 7) | 2.0 (1, 4) | 2.0 (1, 6) |
| Duration^d^ (days), median (min, max) | 2.0 (1, 7) | 1.0 (1, 7) | 2.0 (1, 7) | 2.0 (1, 7) | 2.0 (1, 7) |
| Persisted beyond 7 days | 1 (1.7) | 1 (1.7) | 2 (3.2) | 0 | 0 |

AR, adverse reaction.

^a^Data presented are those from the 2- and 3-dose groups combined (ie, total mRNA-1647 group).

^b^Number of exposed participants who submitted any data for the event.

^c^Percentages are based on the number of exposed participants who submitted any data for the event.

^d^Duration is calculated as the last day - the first day + 1 when the solicited AR was reported starting within the 7 days of injection.

**Supplementary Table 4. Unsolicited TEAE up to 28 days after vaccination regardless of relationship to treatment^a^ (safety set)**

|  | **mRNA-1273.222  (n=60)** | **mRNA-1345  (n=61)** | **mRNA-1647  2-Dose  (n=31)** | **mRNA-1647  3-Dose  (n=31)** | **Total mRNA-1647  (n=62)** | **mRNA-1010  (n=60)** | **FLUAD  (n=59)** | **Total  (N=302)** |
| --- | --- | --- | --- | --- | --- | --- | --- | --- |
| Unsolicited TEAEs regardless of relationship to study vaccination, n (%)^b^ |  |  |  |  |  |  |  |  |
| All | 11 (18.3) | 8 (13.1) | 7 (22.6) | 18 (58.1) | 25 (40.3) | 5 (8.3) | 15 (25.4) | 64 (21.2) |
| Serious | 0 | 1 (1.6) | 0 | 0 | 0 | 0 | 2 (3.4) | 3 (1.0) |
| Fatal | 0 | 0 | 0 | 0 | 0 | 0 | 0 | 0 |
| Medically attended | 2 (3.3) | 4 (6.6) | 5 (16.1) | 9 (29.0) | 14 (22.6) | 1 (1.7) | 8 (13.6) | 29 (9.6) |
| Leading to discontinuation from study   vaccine | 0 | 0 | 0 | 0 | 0 | 0 | 0 | 0 |
| Leading to study discontinuation | 0 | 0 | 0 | 0 | 0 | 0 | 0 | 0 |
| Severe | 0 | 1 (1.6) | 1 (3.2) | 0 | 1 (1.6) | 0 | 2 (3.4) | 4 (1.3) |
| Grade 3 or higher | 0 | 1 (1.6) | 1 (3.2) | 0 | 1 (1.6) | 0 | 2 (3.4) | 4 (1.3) |
| Non-serious | 11 (18.3) | 7 (11.5) | 7 (22.6) | 18 (58.1) | 25 (40.3) | 5 (8.3) | 13 (22.0) | 61 (20.2) |
| Grade 3 or higher non-serious AE | 0 | 0 | 1 (3.2) | 0 | 1 (1.6) | 0 | 0 | 1 (0.3) |
| Occurred in >1 participant in a given vaccine group (any grade), n (%)^b^ |  |  |  |  |  |  |  |  |
| Upper respiratory tract infection | 1 (1.7) | 3 (4.9) | 2 (6.5) | 0 | 2 (3.2) | 1 (1.7) | 2 (3.4) | 9 (3.0) |
| Headache | 2 (3.3) | 2 (3.3) | 1 (3.2) | 3 (9.7) | 4 (6.5) | 0 | 0 | 8 (2.6) |
| Myalgia | 1 (1.7) | 0 | 0 | 3 (9.7) | 3 (4.8) | 0 | 0 | 4 (1.3) |
| Injection site pruritus | 0 | 0 | 1 (3.2) | 2 (6.5) | 3 (4.8) | 0 | 0 | 3 (1.0) |
| Injection site erythema | 0 | 0 | 1 (3.2) | 1 (3.2) | 2 (3.2) | 0 | 0 | 2 (0.7) |
| Injection site induration | 0 | 0 | 0 | 2 (6.5) | 2 (3.2) | 0 | 0 | 2 (0.7) |
| Injection site lymphadenopathy | 0 | 0 | 1 (3.2) | 1 (3.2) | 2 (3.2) | 0 | 0 | 2 (0.7) |
| Oropharyngeal pain | 2 (3.3) | 0 | 0 | 0 | 0 | 0 | 0 | 2 (0.7) |
| Presyncope | 0 | 0 | 0 | 2 (6.5) | 2 (3.2) | 0 | 0 | 2 (0.7) |

AE, adverse event; TEAE, treatment-emergent adverse event.

For each section, subjects who did not report any serious AE were included in the summary of non-serious AEs and grade 3 or higher non-serious AEs. If reported term for the AE contains DEATH or FATAL or outcome of AE contains Fatal, then the toxicity grade is grade 5.

^a^A TEAE is defined as any event not present before exposure to study vaccination or any event already present that worsens in intensity or frequency after exposure.

^b^Percentages are based on the number of subjects in the safety set.

**Supplementary Table 5. Unsolicited treatment-related TEAE up to 28 days after vaccination^a^ (safety set)**

|  | **mRNA-1273.222  (n=60)** | **mRNA-1345  (n=61)** | **mRNA-1647  2-Dose  (n=31)** | **mRNA-1647  3-Dose  (n=31)** | **Total mRNA-1647  (n=62)** | **mRNA-1010  (n=60)** | **FLUAD  (n=59)** | **Total  (N=302)** |
| --- | --- | --- | --- | --- | --- | --- | --- | --- |
| Unsolicited TEAEs related to study vaccination^b^ |  |  |  |  |  |  |  |  |
| All | 4 (6.7) | 1 (1.6) | 4 (12.9) | 11 (35.5) | 15 (24.2) | 0 | 1 (1.7) | 21 (7.0) |
| Serious | 0 | 0 | 0 | 0 | 0 | 0 | 0 | 0 |
| Fatal | 0 | 0 | 0 | 0 | 0 | 0 | 0 | 0 |
| Medically attended | 2 (3.3) | 1 (1.6) | 2 (6.5) | 1 (3.2) | 3 (4.8) | 0 | 0 | 6 (2.0) |
| Leading to discontinuation from study vaccine | 0 | 0 | 0 | 0 | 0 | 0 | 0 | 0 |
| Leading to study discontinuation | 0 | 0 | 0 | 0 | 0 | 0 | 0 | 0 |
| Severe | 0 | 0 | 0 | 0 | 0 | 0 | 0 | 0 |
| Grade 3 or higher | 0 | 0 | 0 | 0 | 0 | 0 | 0 | 0 |
| Non-serious | 4 (6.7) | 1 (1.6) | 4 (12.9) | 11 (35.5) | 15 (24.2) | 0 | 1 (1.7) | 21 (7.0) |
| Grade 3 or higher non-serious AE | 0 | 0 | 0 | 0 | 0 | 0 | 0 | 0 |

AE, adverse event; TEAE, treatment-emergent adverse event.

For each section, subjects who did not report any serious AEs were included in the summary of non-serious AEs and grade 3 or higher non-serious AEs. If reported term for the AE contains DEATH or FATAL or outcome of AE contains Fatal, then the toxicity grade is grade 5.

^a^A TEAE is defined as any event not present before exposure to study vaccination or any event already present that worsens in intensity or frequency after exposure.

^b^Percentages are based on the number of subjects in the safety set.

**Supplementary Table 6. nAb responses after mRNA-1273.222 by baseline SARS-CoV-2 serostatus (per-protocol immunogenicity set)**

|  | **VAC62 nAb (AU/mL)**  **(LLOQ: 10, ULOQ: 111433)** | | | **VAC122 nAb against B.1.1.529 (AU/mL)**  **(LLOQ: 8, ULOQ: 41984)** | | | **VAC137 nAb against BA.4/BA.5 (AU/mL)**  **(LLOQ: 103, ULOQ: 28571)** | | |
| --- | --- | --- | --- | --- | --- | --- | --- | --- | --- |
|  | **SARS-CoV-2 Seropositive**  **(n=48)^a^** | **SARS-CoV-2 Seronegative**  **(n=11)^a^** | **Total**  **(n=60)^a^** | **SARS-CoV-2 Seropositive**  **(n=48)^a^** | **SARS-CoV-2 Seronegative**  **(n=11)^a^** | **Total**  **(n=60)^a^** | **SARS-CoV-2 Seropositive**  **(n=48)^a^** | **SARS-CoV-2 Seronegative**  **(n=11)^a^** | **Total**  **(n=60)^a^** |
| Baseline, n | 48 | 11 | 59 | 48 | 11 | 59 | 48 | 11 | 59 |
| GMC (95% CI) | 2994.29  (2306.02, 3887.98) | 480.00 (184.34, 1249.84) | 2128.45 (1543.15, 2935.74) | 581.91 (418.87, 808.40) | 60.71  (27.10, 136.02) | 381.81 (262.30, 555.77) | 286.78 (201.04, 409.09) | 62.62 (46.23, 84.81) | 215.94 (155.22, 300.42) |
| Day 29, n | 47 | 11 | 58 | 47 | 11 | 58 | 47 | 11 | 58 |
| GMC (95% CI) | 10869.86 (8955.42,  13259.19) | 7165.60 (4661.38,  11015.16) | 10086.00 (8410.68,  12095.01) | 3289.08 (2592.73, 4172.45) | 1400.73 (792.06, 2477.13) | 2789.13 (2258.74, 3444.06) | 1813.48 (1433.73,  2293.82) | 690.38 (429.32,  1110.19) | 1507.59 (1228.20,  1850.53) |
| GMFR (95% CI) | 3.63 (2.98, 4.42) | 14.84 (9.65, 22.81) | 4.70 (3.92, 5.64) | 5.62 (4.43, 7.13) | 23.07 (13.05, 40.80) | 7.29 (5.90, 9.00) | 6.33 (5.01, 8.01) | 11.03 (6.86, 17.73) | 7.00 (5.70, 8.59) |
| SRR (95% CI), % ^b^ | 42.6  (28.26, 57.82) | 63.6  (30.79, 89.07) | 46.6  (33.34, 60.13) | 57.4  (42.18, 71.74) | 90.9  (58.72, 99.77) | 63.8 (50.12, 76.01) | 55.3 (40.12, 69.83) | 72.7  (39.03, 93.98) | 58.6  (44.93, 71.40) |

CI, confidence interval; GMFR, geometric mean fold rise; GMC, geometric mean concentration; LLOQ, lower limit of quantification; nAb, neutralizing antibody; SRR, seroresponse rate; ULOQ, upper limit of quantification.

Antibody values <LLOQ or >ULOQ were replaced by 0.5 x LLOQ or converted to the ULOQ, respectively.

^a^Number of participants with non-missing SARS-CoV-2 serology sample test results at baseline.

^b^SRR was defined as a post-vaccination titer ≥4-fold x LLOQ if the baseline titer was <LLOQ or as a post-vaccination titer ≥4-fold of the baseline titer if ≥LLOQ.

**Supplementary Table 7. RSV-A and RSV-B nAb responses after mRNA-1345 by age-group (per-protocol immunogenicity set)**

|  | **Age group** | | | | | **Total**  **n=60^a^** | | |
| --- | --- | --- | --- | --- | --- | --- | --- | --- |
|  | **18-49 years**  **n=31^a^** | | **50-75 years**  **n=29^a^** | | |  |  |  |
|  | **RSV-A** | **RSV-B** | | **RSV-A** | **RSV-B** | | **RSV-A** | **RSV-B** |
| Baseline, n | 31 | 31 | | 29 | 29 | | 60 | 60 |
| GMT (95% CI) | 1502.48 (1116.64, 2021.65) | 1281.24 (914.99, 1794.09) | | 2362.96 (1365.14, 4090.13) | 1525.57 (1094.72, 2126.00) | | 1870.06 (1381.74, 2530.94) | 1394.02 (1106.85, 1755.69) |
| Day 29, n | 27 | 27 | | 29 | 29 | | 56 | 56 |
| GMT (95% CI) | 15325.27 (11003.19, 21345.07) | 6326.10 (4534.32, 8825.91) | | 14842.46 (10249.70, 21493.17) | 6224.73 (4676.45, 8285.61) | | 15122.64 (11915.27, 19193.39) | 6236.02 (5077.95, 7658.18) |
| GMFR (95% CI) | 10.09 (7.25, 14.06) | 4.98 (3.57, 6.95) | | 6.28 (4.34, 9.10) | 4.08 (3.07, 5.43) | | 7.98 (6.29, 10.13) | 4.48 (3.65, 5.50) |
| SRR (95% CI), %^b^ | 85.2 (66.27, 95.81) | 51.9 (31.95, 71.33) | | 58.6 (38.94, 76.48) | 55.2 (35.69, 73.55) | | 71.4 (57.79, 82.70) | 53.6 (39.74, 67.01) |

CI, confidence interval; GMFR, geometric mean fold rise; GMT, geometric mean titer; LLOQ, lower limit of quantification; nAb, neutralizing antibody; RSV, respiratory syncytial virus; SRR, seroresponse rate; ULOQ, upper limit of quantification.

Antibody values <LLOQ or >ULOQ were replaced by 0.5 x LLOQ or converted to the ULOQ, respectively.

^a^Number of participants with non-missing nAb data at baseline and on Day 29.

^b^SRR was defined as a post-vaccination titer ≥4-fold x LLOQ if the baseline titer was <LLOQ or as a post-vaccination titer ≥4-fold of the baseline titer if ≥LLOQ.

**Supplementary Table 8. Anti-HA antibody responses after mRNA-1010 and FLUAD (per-protocol immunogenicity set)**

|  | **mRNA-1010** | | | | **FLUAD** | | | |
| --- | --- | --- | --- | --- | --- | --- | --- | --- |
|  | **A/H1N1** | **A/H3N2** | **B/Victoria** | **B/Yamagata** | **A/H1N1** | **A/H3N2** | **B/Victoria** | **B/Yamagata** |
| Baseline, n^a^ | 55 | 55 | 55 | 55 | 57 | 57 | 57 | 57 |
| GMT (95% CI) | 41.00 (32.27, 52.09) | 27.38 (22.37, 33.51) | 65.85 (53.71, 80.73) | 57.29  (44.33, 74.02) | 41.44  (32.29, 53.20) | 33.30 (26.69, 41.56) | 65.08  (54.60, 77.57) | 56.52 (42.07, 75.92) |
| Day 29, n | 55 | 55 | 55 | 55 | 57 | 57 | 57 | 57 |
| GMT (95% CI) | 201.12 (154.23,262.15) | 112.19 (87.95,143.10) | 100.01 (82.28,121.55) | 149.58 (121.38,184.34) | 241.47 (186.12,313.28) | 136.88  (107.78, 173.85) | 129.09  (106.57, 156.35) | 149.34  (121.63, 183.37) |
| GMFR (95% CI) | 4.88 (3.74,6.36) | 3.71 (2.91,4.73) | 1.53 (1.26, 1.86) | 2.63 (2.13, 3.24) | 5.86 (4.51, 7.60) | 4.53 (3.56, 5.75) | 1.97 (1.63, 2.39) | 2.63 (2.14, 3.22) |
| SRR (95% CI), %^b^ | 67.3 (53.29,79.32) | 54.5  (40.55, 68.03) | 9.1  (3.02, 19.95) | 34.5  (22.24, 48.58) | 61.4  (47.57, 74.00) | 59.6 (45.82,72.44) | 24.6  (14.13, 37.76) | 35.1  (22.91, 48.87) |

A/H1N1, A/Wisconsin/588/2019 (H1N1) pdm09-like virus; A/H3N2, A/Darwin/6/2021 (H3N2)-like virus; B/Victoria, B/Austria/1359417/2021

(B/Victoria lineage)-like virus, B/Yamagata, B/Phuket/3073/2013 (B/Yamagata lineage)-like virus; CI, confidence interval; GMFR, geometric mean fold rise; GMT, geometric mean titer; HA, hemagglutinin; LLOQ, lower limit of quantification; SRR, seroresponse rate; ULOQ, upper limit of quantification.

Antibody values <LLOQ or >ULOQ were replaced by 0.5 x LLOQ or converted to the ULOQ, respectively.

^a^Number of participants with non-missing anti-HA results at baseline and day 29

^b^SRR was defined as a post-vaccination titer ≥4-fold x LLOQ if the baseline titer was <LLOQ or as a post-vaccination titer ≥4-fold of the baseline titer if ≥LLOQ.
